# Artificial Intelligence in Cardiac Surgery: A Systematic Review

**DOI:** 10.1101/2023.10.18.23297244

**Authors:** Ralf Martz Sulague, Francis Joshua Beloy, Jillian Reeze Medina, Edward Daniel Mortalla, Thea Danielle Cartojano, Sharina Macapagal, Jacques Kpodonu

**Author notes:** Corresponding Author: Jacques Kpodonu, MD Division of Cardiac Surgery, Department of Surgery, Beth Israel Deaconess Medical Center 330 Brookline Ave, Boston, MA 02215.

## Abstract

**BACKGROUND:** Artificial intelligence has emerged as a tool to potentially increase efficiency and efficacy of cardiovascular care and improve clinical outcomes. This study aims to provide an overview of applications of artificial intelligence in cardiac surgery.

**METHODS:** A systematic literature search on artificial intelligence applications in cardiac surgery from inception to February 2024 was conducted. Articles were then filtered based on the inclusion and exclusion criteria and risk of bias was assessed. Key findings were then summarized

**RESULTS:** A total of 81 studies were found that reported on artificial intelligence applications in cardiac surgery. There is a rapid rise in studies since 2020. The most popular machine learning technique was Random Forest (n=48), followed by Support Vector Machine (n=33), Logistic Regression (n=32), and Extreme Gradient Boosting (n=31). Most of the studies were on adult patients, conducted in China, and involved procedures such as valvular surgery (24.7%), heart transplant (9.4%), coronary revascularization (11.8%), congenital heart disease surgery (3.5%), and aortic dissection repair (2.4%). Regarding evaluation outcomes, 35 studies examined the performance, 26 studies examined clinician outcomes, and 20 studies examined patient outcomes.

**CONCLUSION:** Artificial intelligence was mainly used to predict complications following cardiac surgeries and improve clinicians’ decision-making by providing better preoperative risk assessment, stratification, and prognostication. While the application of artificial intelligence in cardiac surgery has greatly progressed in the last decade, further studies need to be done to verify accuracy and ensure safety before use in clinical practice.

## INTRODUCTION

With the advancement of modern technology, artificial intelligence (AI) has emerged as a tool to potentially increase the efficiency and efficacy of healthcare and improve outcomes. It encompasses both machine learning (ML) and deep learning (DL). In ML, certain computer algorithms are used to produce predictions or conclusions by recognizing patterns generated through the application of a mathematical algorithm model from sample data. One important example of the significance of machine learning in surgery would be predicting the probabilities of post-operative complications according to patient specific risk factors and characteristics. It would use the data to classify patients into risk strata, depending on their morbidity severity. It is able to do so with great accuracy, exceeding previous methods based on clinical standards to levels previously thought to be unachievable with conventional statistics. On the other hand, DL uses a multi-layered structure of algorithms called artificial neural networks to do tasks that machine learning cannot, making it more useful than machine learning (ML) (1,2).

There have been a number of studies exploring real-life applications of AI in cardiac surgery including algorithms that function to aid in clinical decision-making, especially in terms of cardiac function evaluation and risk stratification prior to operation. Other applications focus on aiding diagnostics and prognostication of certain complications of patients after cardiac surgery(3).

The growing body of knowledge of AI applications in cardiac surgery necessitates evaluation of past studies to gain insights to the future direction of artificial intelligence application in cardiac surgery. This study aims to provide an overview of the applications of AI in cardiac surgery through a systematic review.

## METHODS

### Search Strategies

The Preferred Reporting Items for Systematic Reviews and Meta-Analyses (PRISMA) guidelines were utilized in searching articles assessing and evaluating various applications of AI in cardiac surgery from inception to February 2024. Using boolean search terms “Artificial Intelligence” OR “Machine Learning” AND “Cardiac Surgery”, a thorough review of studies was conducted using the following databases: PubMed, Embase, Europe PMC, Epistemonikos, CINAHL, Cochrane Central, Google Scholar, Web of Science, Scopus, Cambridge Core, clinicaltrials.gov, and science.gov. Duplicate articles from different databases were then excluded after a preliminary search. Other additional studies were identified by looking through the references of the articles that were already included. This systematic review was registered on Prospero (CRD42022377530).

### Eligibility Criteria

Articles were incorporated into the review if it included the following conditions: 1) Implementation of an AI application with patient or health care providers in a real-life clinical setting, and 2) Provision of decision support by the AI application through emulating clinical decision-making processes of health care providers (eg, medical image interpretation and clinical risk assessment). All cohort studies and randomized control trials on adult cardiac surgery that satisfied the inclusion criteria were included. The studies that were included had to be in English. Studies that had only been published as abstracts, review papers, meta-analyses, clinical trials that were still in progress, and published study protocols were not included. Other exclusion criteria are detailed in Figure 1.

**FIGURE 1.**
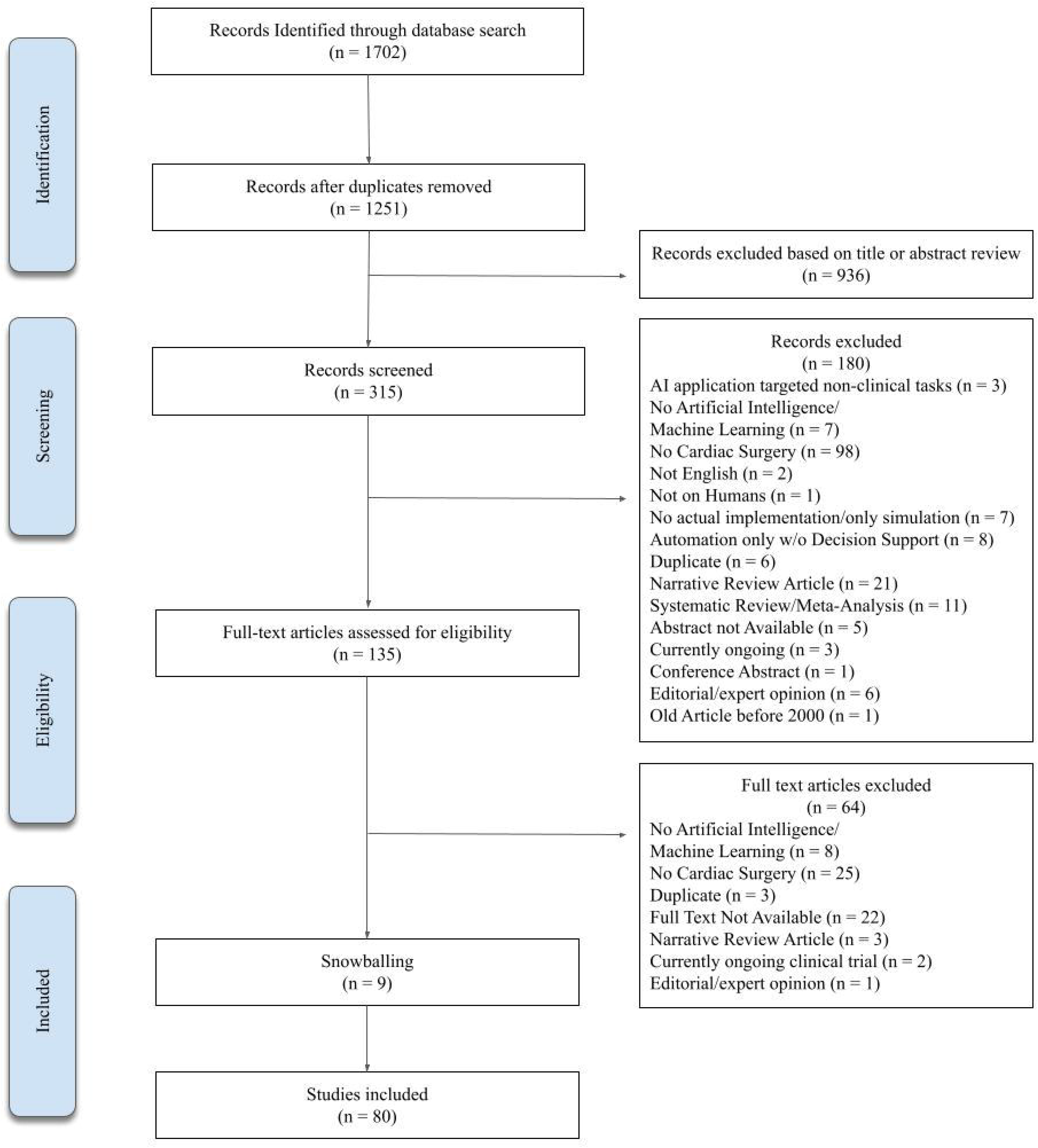
Preferred Reporting Items for Systematic Reviews and Meta-analyses (PRISMA) flow diagram.

### Data Extraction

After collating all the studies, information from the articles were extracted. These were primarily the characteristics of the studies, the features of the AI applications, and the key outcomes evaluated. The data was then organized in a table and trends or themes were analyzed.

## RESULTS

### Overview

Our initial search of the databases returned a total of 1702 journal articles (617 from PubMed, 190 from Google Scholar, 517 from Web of Science, 165 CINAHL, 130 from EMBASE, 69 from Europe PMC, 6 from Epistemonikos, 3 from science.gov, 2 from clinicaltrials.gov, 2 from Cambridge Core, and 1 from Cochrane Central). Duplicates were then identified and 451 studies were excluded. The titles, abstracts, and manuscripts were screened and filtered which excluded 1179 studies. Additional 9 relevant studies were identified via snowballing. Thus, 81 journal articles met the inclusion criteria and were included in this review (Figure 1).

### Study Characteristics

The authors, year of publication, study design, number of involved patients, and database registry and involved hospitals were summarized in Table 1.

**Table 1.**
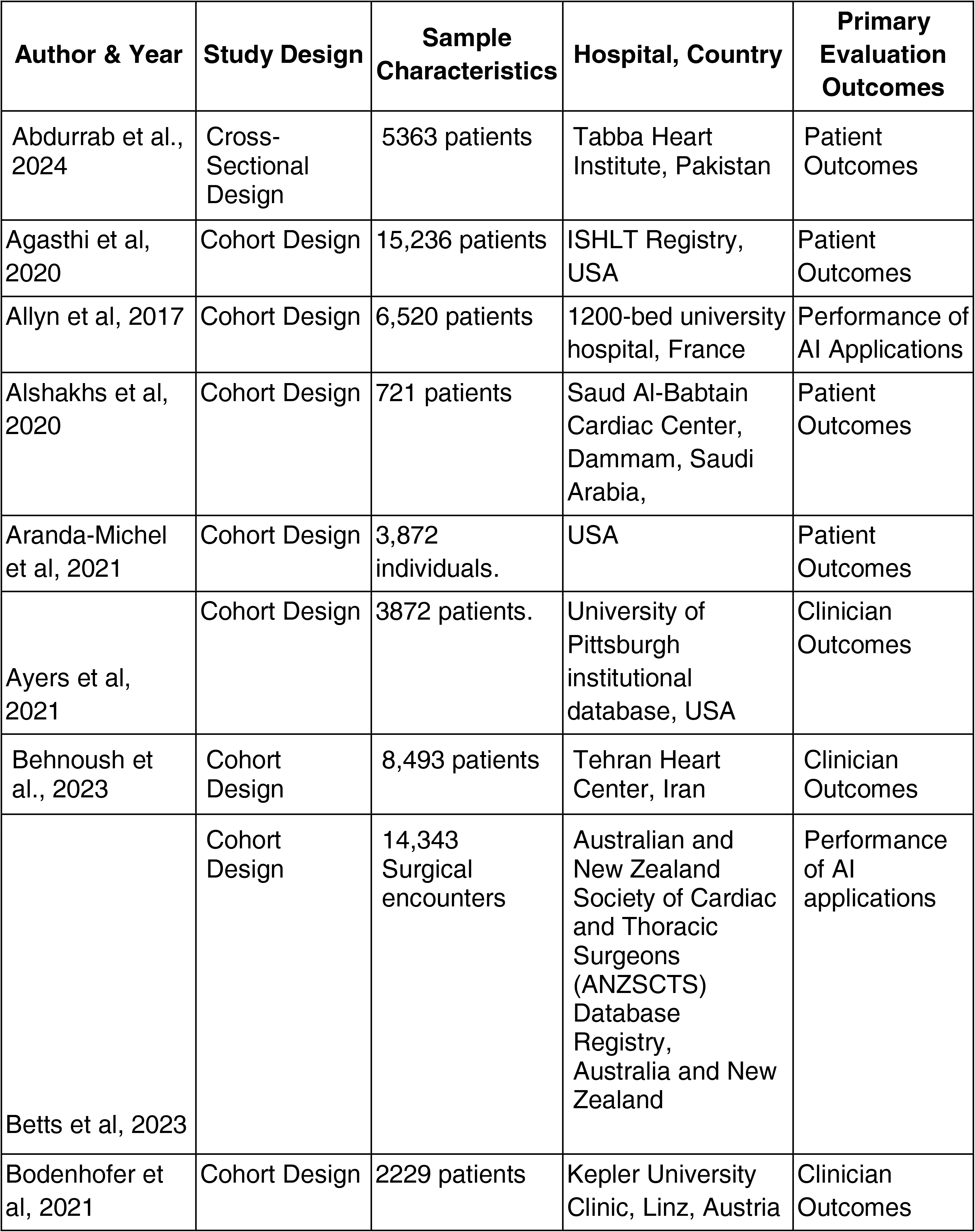
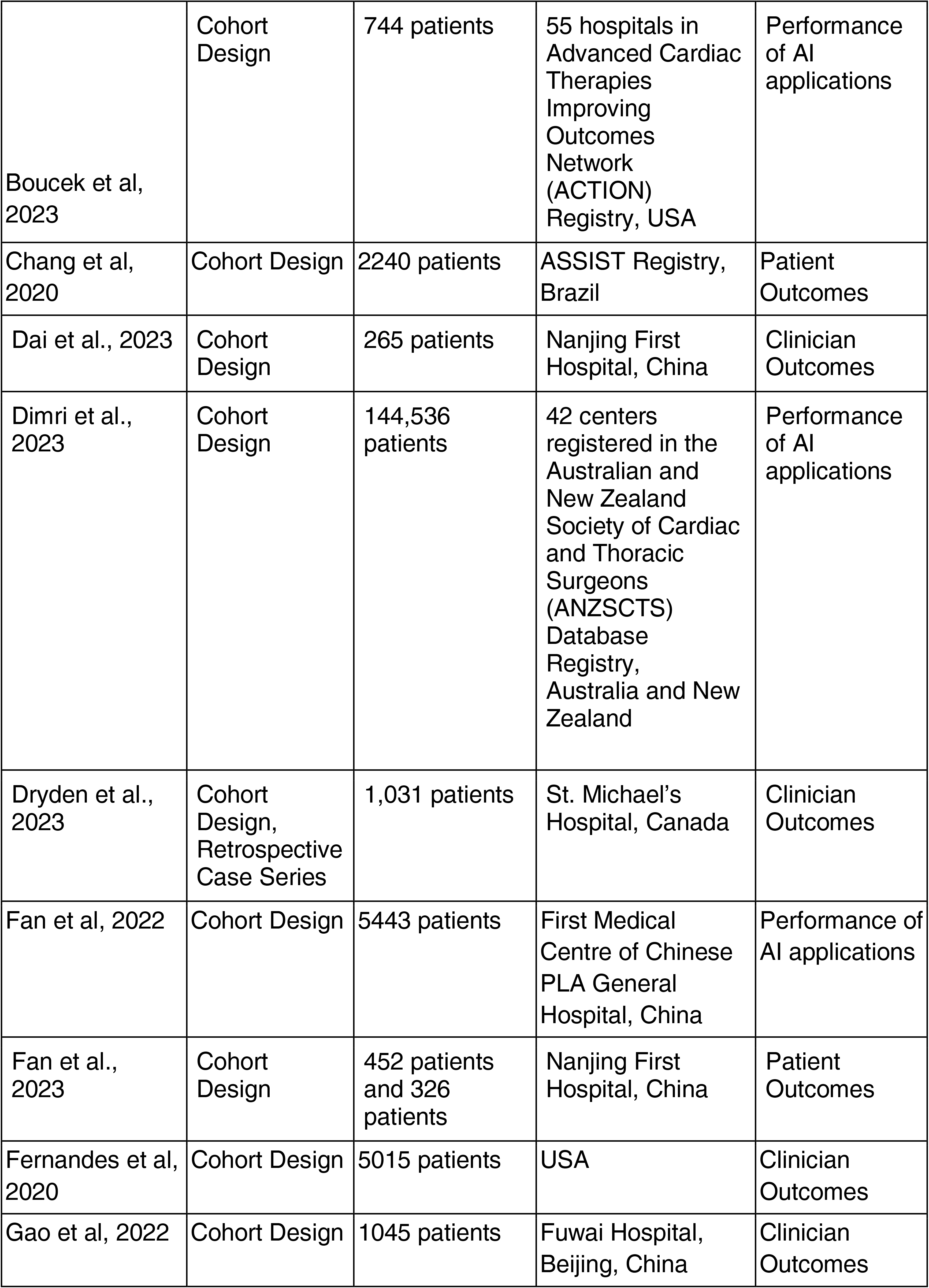
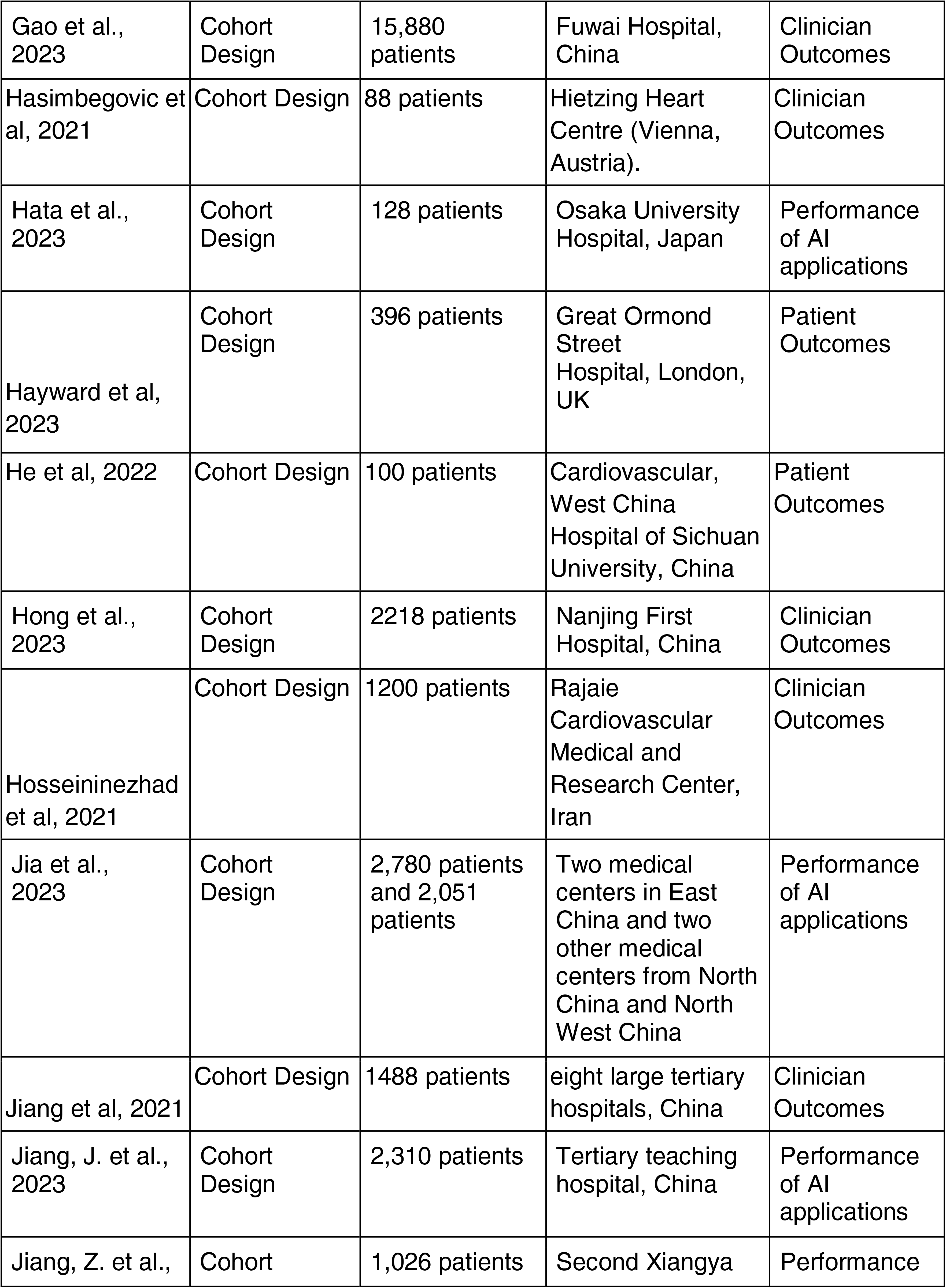
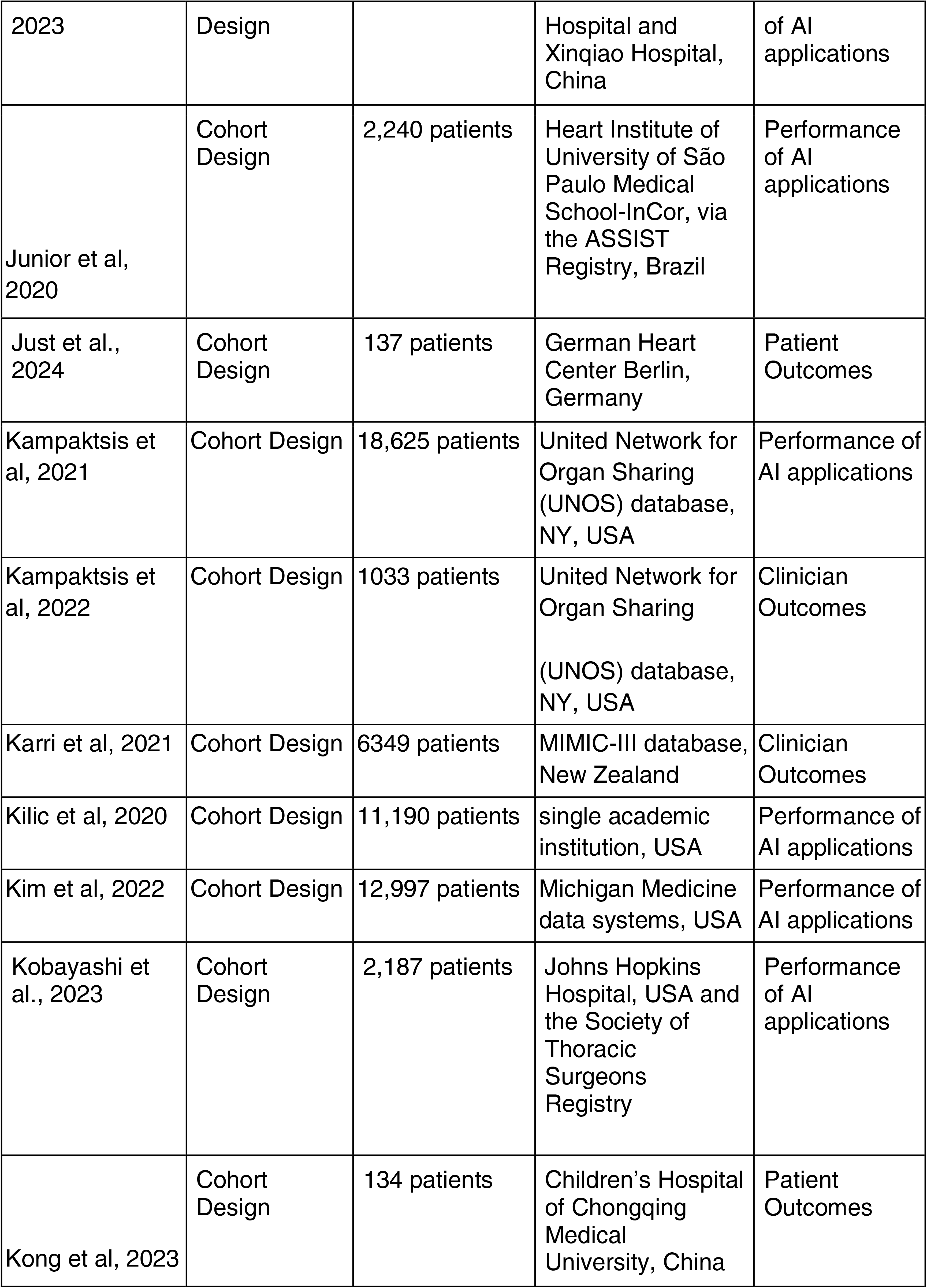
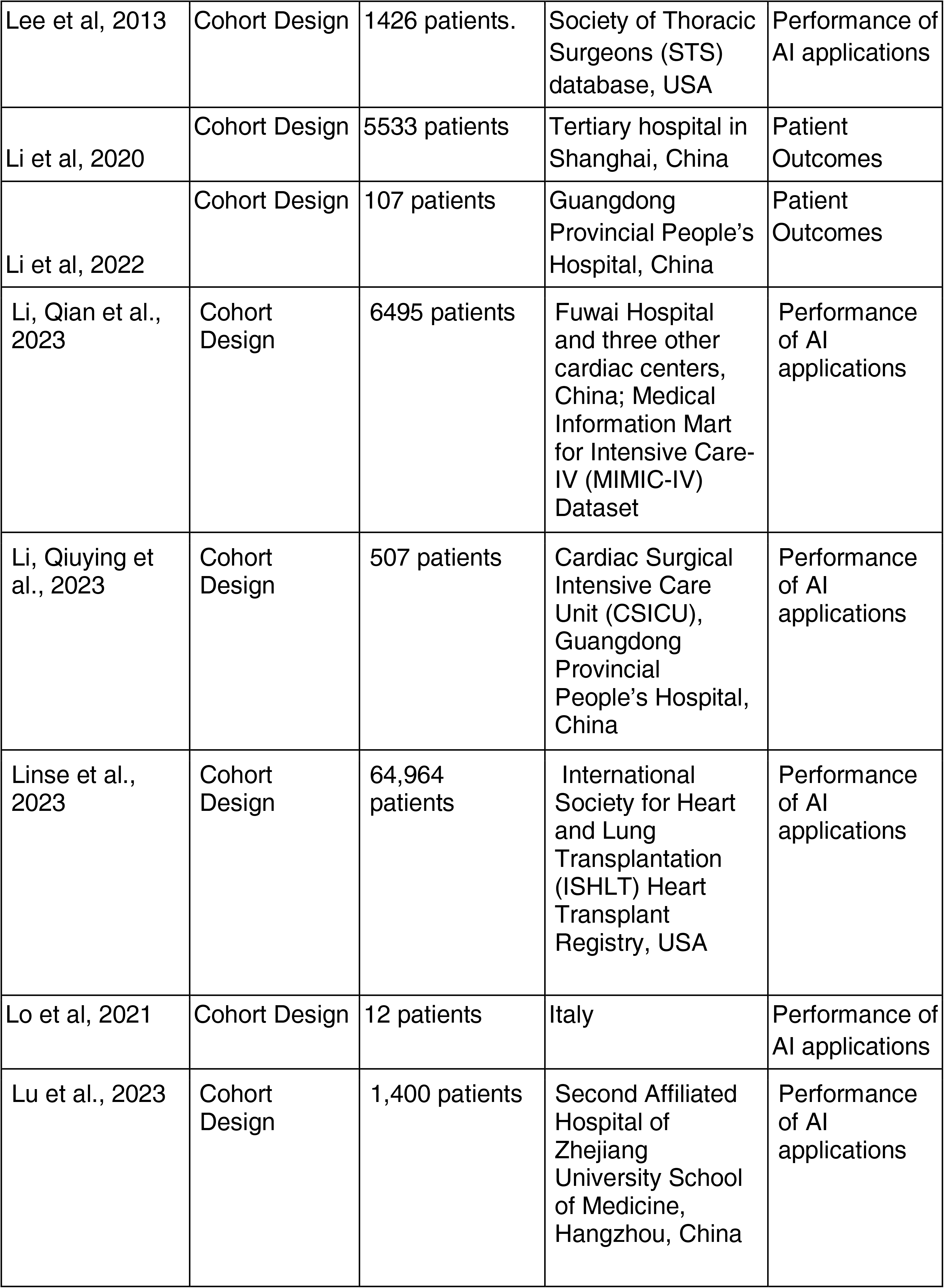
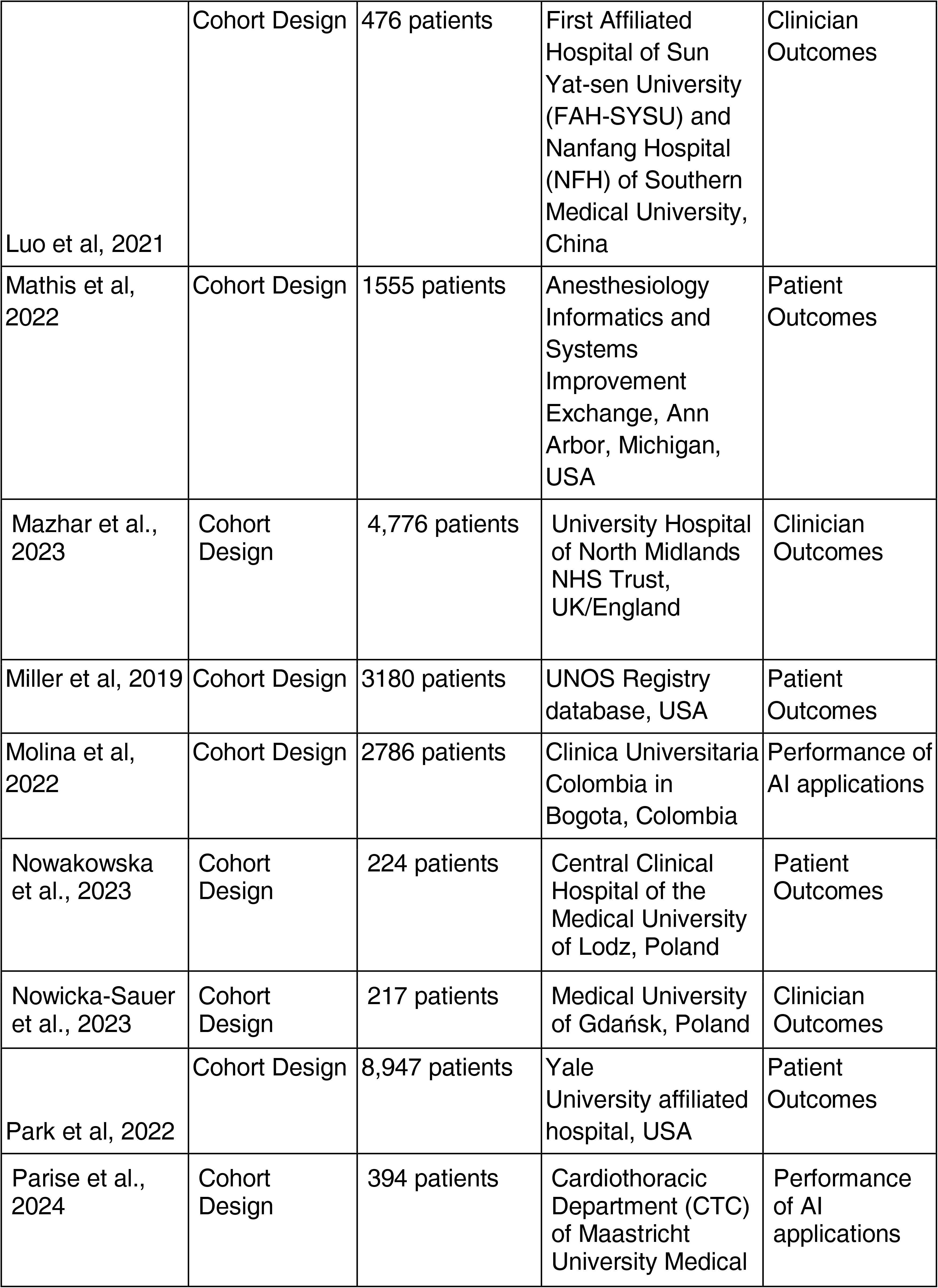
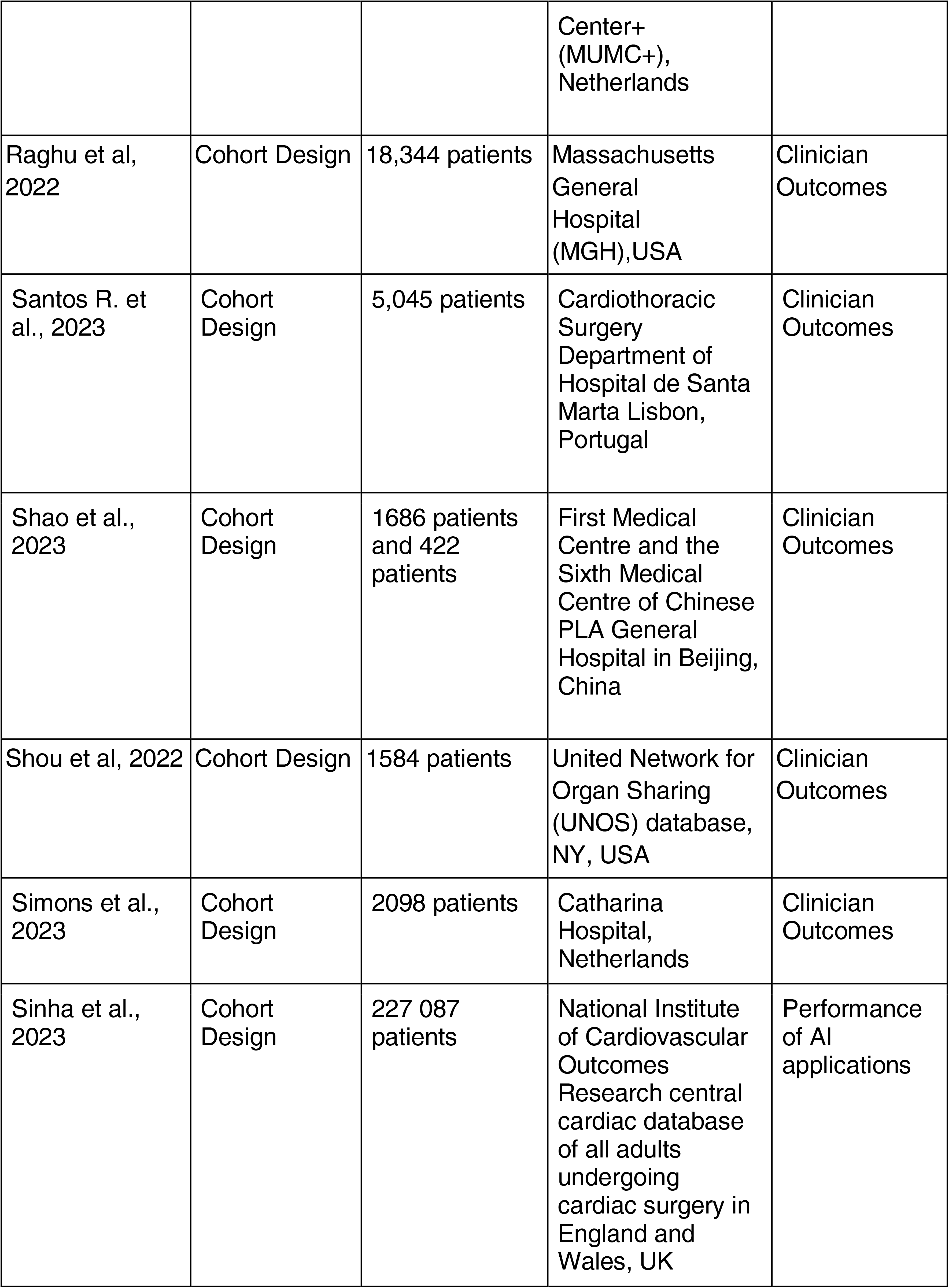
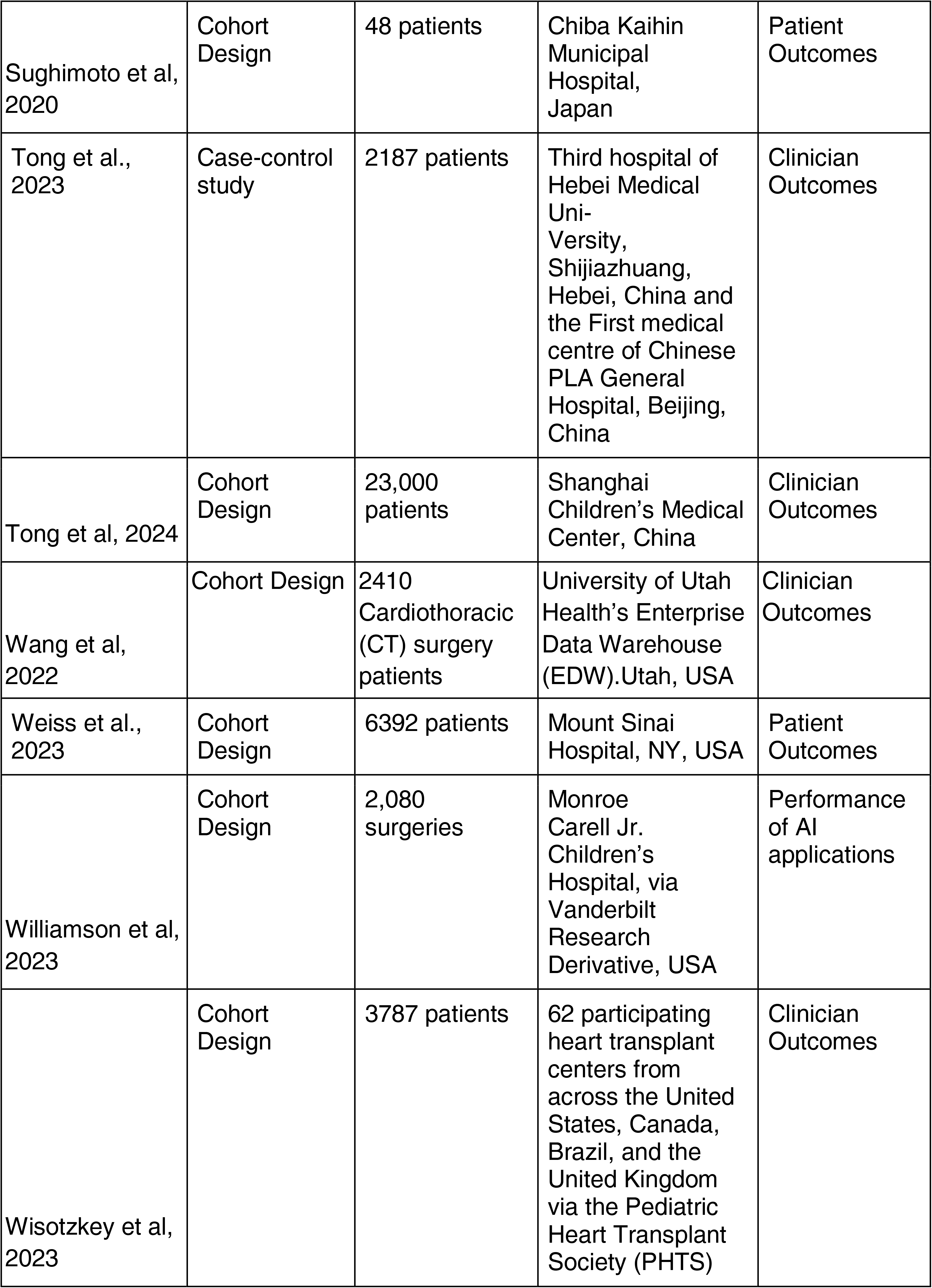
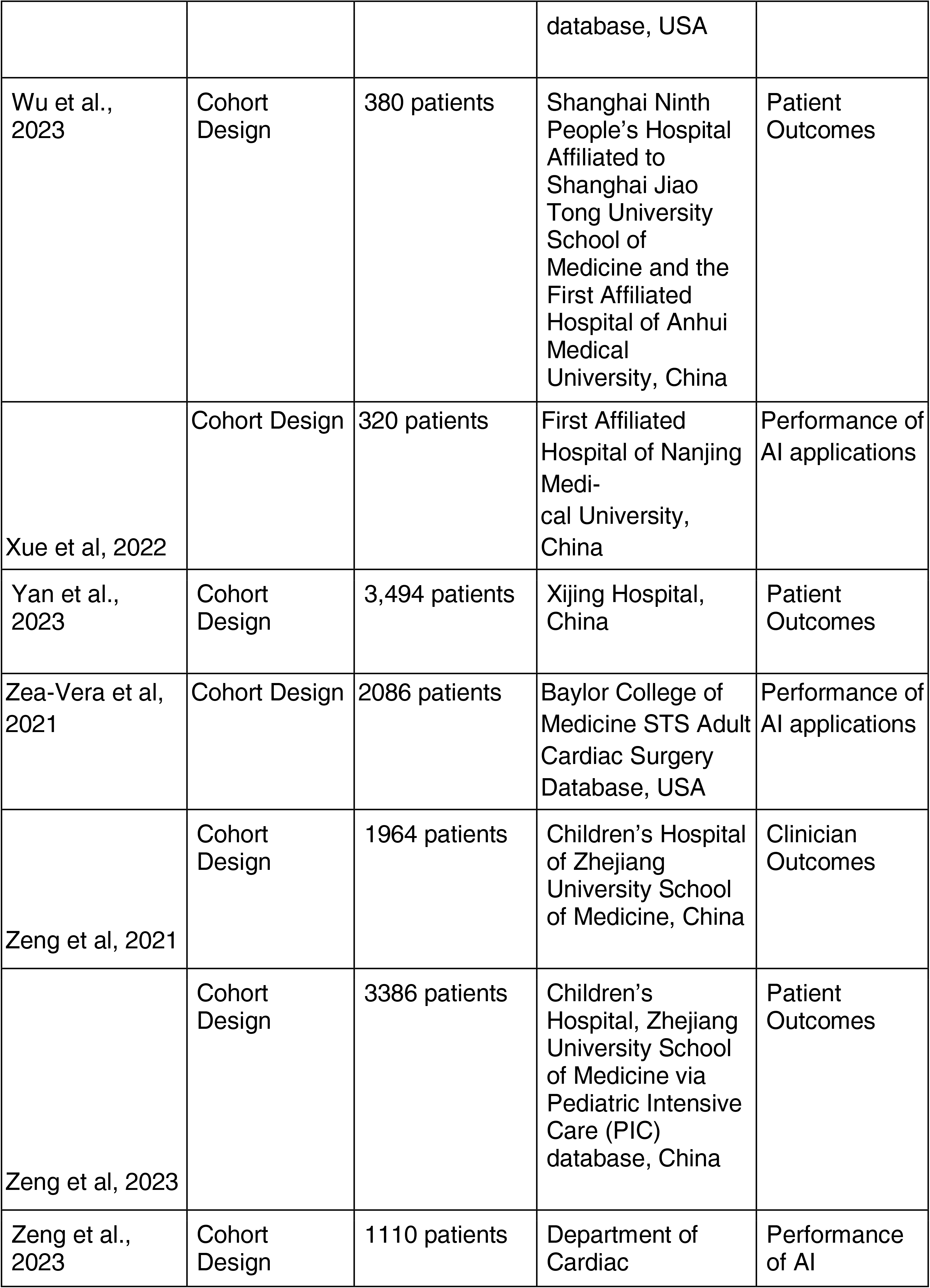
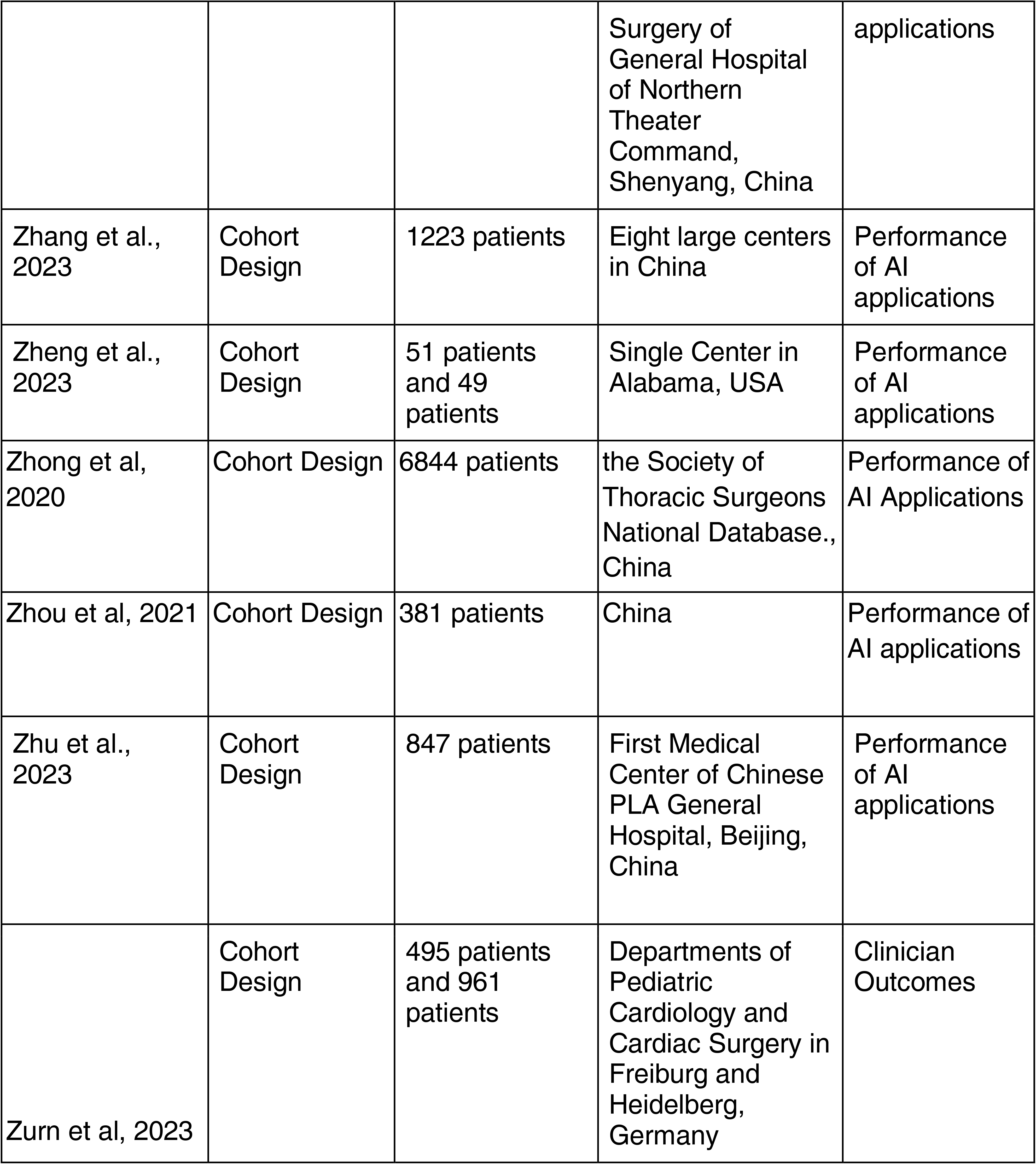
Characteristics of the studies included.

Figure 2 highlights an increasing trend in the number of published studies on the application of artificial intelligence in cardiac surgery in the past ten years, with an observed rapid rise since 2020 suggesting increased interest in the field in recent years. Although the systematic review included studies until February 2024, this timeline was not included in the graph since the year has not ended and will only add a false decrease in trend when instead, it is anticipated to increase even more.

Majority of the studies were cohort studies. One study made use of a cross-sectional study design, another utilized a case-control design and one other study utilized a combination of a cohort and retrospective case series. Of the 81 studies, two (2.5%) studies have less than 50 patients analyzed. Three (3.7%) studies have a population of 50-100 patients, while 34 (42.0%) studies have a wider range of 1000- 5000 individuals included. Meanwhile, 19 (23.5%) studies had a larger scaled population of 5,000-20,000 patients, and one (1.2%) study had more than 220,000 worth of patient data analyzed.

Figure 3 showed the geographic distribution of the published studies based on where the study was conducted. About 12 (28.57%) studies mentioned the involved hospitals or clinics while 6 (13.29%) of these did not specify. About 17 (40.48%) studies utilized a database registry for their data, while 7 (16.67%) had no mention of their population groups.

Of the 81 studies, China, an upper-middle income country, had the most contribution with 31 studies (Figure 3) while only 2 studies were notably conducted in a lower-middle income countries, namely Iran and Pakistan. For other countries classified Upper-Middle Income Economies, Colombia and Brazil had conducted 1 and 2 studies, respectively. The rest were conducted in countries with high-income economies, with 23 conducted in the United States, two in Austria and Germany, respectively, and one each in Saudi Arabia, Canada, Italy, and France. The country classification was based on the New World Bank country classifications by income level: 2022-2023, for the current 2023 fiscal year, low-income economies are defined as those with a GNI per capita, calculated using the World Bank Atlas method, of $1,085 or less in 2021; lower middle-income economies are those with a GNI per capita between $1,086 and $4,255; upper middle-income economies are those with a GNI per capita between $4,256 and $13,205; high-income economies are those with a GNI per capita of $13,205 or more.

### Quality Assessment

In order to evaluate the internal validity of the 81 included studies, the Joanna Briggs Institute (JBI) critical appraisal tool was utilized. Each article was evaluated using the appropriate checklist by two assessors. The total score for the cohorts ranged from 7 to 10 out of 11. Specifically, four studies reported that the source of their data did not come from the same population (4–7). Seven studies were unable to identify the confounding factors (5,8–13). Only three studies did not clearly state if exposures were measured in a valid and reliable way (4–6). Finally, all studies used proper statistical analysis, and valid outcome measurement methodology.

### AI Application Characteristics

Majority of the studies were conducted on adult cardiac patients 68 (84%) while some were on pediatric cardiac patients 13 (17%) (Figure 4A). Although most of the studies 43 (53%) did not have a specific surgical procedure involved (Figure 4B), 12 (14.81%) studies focused on Coronary Artery Bypass Grafting (CABG), 10 (12.35%) on Heart Transplantation (HT), 9 (11.11%) on Valvular Surgery, 4 (4.94%) on Congenital Heart Surgery, 2 (2.47%) on Ventricular Assist Device (VAD) implantation, and 1 (1.23%) on Aortic Dissection Surgery.

The results also show that almost all AI applications provided decision support in Risk Analysis (n=40) mainly predicting mortality outcomes, post-operative complications, or post-operative outcomes. Only two studies looked into disease screening and triage.

Among the 81 studies (Figure 5, Supplemental Table 1), the most popular ML technique was Random Forest (RF) (n=48), followed by Support Vector Machine (n=33), Logistic Regression (LR) (n=32), and Extreme Gradient Boosting (XGBoost) (n=31). These were followed by AdaBoost, Decision Tree (DT), and K-nearest neighbors classifier (KNN), which were all utilized by 13 (n=13) studies each, respectively. The other applications featured were the following: Gradient Boosting Machine (GBM) (n=11), Multilayer Perceptron (MLP) (n=11), Naïve Bayes Model (NB) (n=10), Artificial Neural Network (ANN) (n=8), Light Gradient Boosting Machine (LGBM) (n=8), Gradient Boosting Decision Trees (GBDT) (n=7), Neural Networks (Nnet) (n=7), CatBoost (n=5), Extra Trees (ET) (n=4), Bag Decision Trees (BDT) (n=3), Bayesian

Networks (BN) (n=3), Gaussian Naive Bayes (GNB) (n=3), Bagged Classification and Regression Tree (CART) (n=2), Cox Regression Models (n=2), Stochastic Gradient Boosting (SGB) (n=2), Lasso Regression (n=2), Support Vector Regression (SVR) (n=2), (Multivariate Adaptive Regression Splines (MARS) (n=2), Deep Neural Network (DNN) (n=2), and Linear Regression (n=2).

The rest of the ML techniques were mentioned only in one study such as the Boosted Classification Trees, Conditional Inference Random Forest (CIRF), Deep Learning Model (CXR-CTSurgery), Dual-tree complex wavelet packet transform (DTCWPT), Gaussian Process (GP) regression ML algorithm, GenAlgs, Imbalanced Random Forest Classifier, Multivariate logistic regression (MLR), Random Forest Survival Model, Stochastic Gradient Boosting (SGBT), Sun Yat-sen University Prediction Model for Infective Endocarditis, Stochastic Gradient Descent Regression, Huber Regression, Ridge Regression, Multiple Linear Regression, Penalized Linear Regression, Deep Forest Model, Softmax Regression, Bootstrapped Aggregation/Bagging, Convolutional Neural Network (CNN), Recurrent Neural Network (RNN), Transformer, Perceptron, Support Vector Machine with Radial Basis Function Kernel, Complement Naive Bayes, AutoML, Linear Discriminant Analysis, Logistic Regression with L2 Regularization, Subspace Discriminant, Subspace KNN, Random Under-Sampling (RUS) Boosted Trees, Logistic Regression with Elastic Net Regularization, Hypertuned RF, RF Regressor, AdaBoost Regressor, Hypertuned AdaBoost, Hypertuned Decision Tree, Logistic Regression with Ridge Penalization, Axis-based Random Survival Forests, Oblique Random Survival Forests, Long Short-Term Memory RNN, Gate Recurrent Units (GRU) RNN, Dipole, RETAIN, and Time Aware Attention RNN.

### Evaluation Outcomes

The included studies were classified into their respective type of evaluation outcomes: performance of AI applications, clinician outcomes, and patient outcomes (see Table 1, summarized in Figure 4C).

#### Performance of AI Applications

Thirty-five studies evaluated the performance of AI applications in real-life clinical settings. Commonly used performance metrics included accuracy, area under the curve (AUC)/area under the receiver operating characteristic curve (AUROC), specificity, sensitivity, True Positive Rate (TPR), False Negative Rate (FNR), positive-predictive value (PPV), and negative-predictive value (NPV), F1 score and Brier Score.

#### Clinician Outcomes

AI applications also affect clinician outcomes, specifically, clinician decision making, clinician workflow and efficiency, and clinician evaluations and acceptance of AI applications. In this review, twenty-six reported clinician outcomes of AI in cardiac surgery (4,5,7,8,10,13–38).

Clinicians could potentially be guided by AI applications in making better medical decisions. Eighteen studies reported that AI applications can support clinician decision making (4,7,10,13,16,17,19,21,23,24,26,28,30–33,35,38). Machine learning models improve clinician’s medical decisions by providing better preoperative risk assessment, stratification and prognostication (10,17,21,24,30–32,35,38). AI applications could also guide clinicians on how aggressive prophylactic measures are given such as increased patient monitoring or giving additional therapies (4,13,33).

Twelve studies discussed clinician efficiency (5,10,16,18,20,22–24,29,36,37). Machine learning was used to predict survival after heart transplantation allowing better patient selection and reducing organ wastage (18,37). AI applications could also prompt clinicians to provide timely protective strategies which will improve patient’s prognosis (1,29,36). AI applications save time significantly by optimizing risk stratification and clinical management. Alshakhs et al. (16) took advantage of machine learning to predict patients who are likely to have a longer postoperative length of stay (PLoS) to provide early psychosocial preparation to the patient and to their family. There were no studies that explored outcomes on clinician workflow.

Seven studies reported clinician evaluations and acceptance of AI applications (5,15,17,19,20,22–24,26). All of the studies stated overall positive perceptions on AI applications. Machine learning showed equal risk prediction compared to manual approaches (20,24). Two studies revealed superiority of AI applications than existing scoring tools (15,17,19,22,26). Finally, recommendations were provided on utilizing both machine learning and manual approach in combination to provide significant leaps in diagnostic and predictive capabilities of clinicians in the future (5,23).

#### Patient Outcomes

Only twenty studies reported patient outcomes. Fernandez *et al*. (21) incorporated intraoperative risk factors in predicting mortality following cardiac surgery and revealed results on patient mortality which revealed the following findings: (1) all deaths, regardless of cause, occurring during the hospitalization in which the operation was performed, even if after 30 days (including patients transferred to other acute care facilities); and (2) all deaths, regardless of cause, occurring after discharge from the hospital, but before the end of the thirtieth postoperative day.

Zea-Vera *et al*. (6) developed and validated a dynamic machine learning model to predict CABG outcomes at clinically relevant pre-and postoperative time points. Their ML predicted 30-day readmission and high cost, 2 outcomes for which no standardized regression model exists. With reduction in mortality, resource utilization is becoming an increasingly important outcome.

## DISCUSSION

### Principal Findings

In recent years, the rise of AI has grown dramatically and transformed how people learn and complete tasks especially in medicine and surgery. ML algorithms have impacted surgical care by assisting the surgeons in making better clinical decisions in the preoperative and intraoperative phases of surgical procedures. These AI applications aim to enhance patient safety by optimizing patient outcomes and surgical decision-making. In this review, we discuss the significant advancements and promising applications of AI in cardiac surgery and its risks. The growing interest in its application to surgical practice produced the following findings.

To provide accurate analysis, we only included English–written articles discussing the actual implementation of AI in real-life clinical settings. The majority of the included papers were published between 2020 and 2022 in order to give the most recent information on the use of AI in cardiac surgery. Most of the reviewed studies utilizes a cohort study design with a database registry composed of 1000-5000 participants per study.

It is worth noting that half of the included studies were from the United States, which suggests that developed countries are in the forefront of AI application in health care. Recently, the AI algorithms were being used in analyzing factors contributing to COVID-19 mortality and detection of pathological findings (2,39). In cardiac surgery, ML algorithms were used to predict mortality, survival, postoperative length of stay, and outcomes in following cardiac surgeries such as valve replacement, coronary artery bypass graft surgery, and heart transplantation (10,12,16,18,38,40). More than half of the studies utilized RF as ML technique to predict mortality and outcomes after cardiac surgery. Aside from the United States, China has been making substantial use of AI in healthcare. In the review, ten studies were conducted in China, largely in specified hospitals. Due to their capacity to generate customized risk profiles, ML models have the potential to show better predictive power for risk stratification compared to clinical scores like EuroSCORE (24).

### Lack of funds

Insufficient funding poses a significant obstacle to the integration of AI in clinical practice, particularly in the field of cardiac surgery. The successful development and deployment of AI systems necessitate substantial financial resources. These include investments in infrastructure, data acquisition and management, algorithm development, and training. Unfortunately, numerous healthcare institutions encounter difficulties in allocating the required funds to support AI initiatives, given competing priorities and limited budgets(41). Insufficient financial support hampers the seamless integration of AI technologies into cardiovascular surgery practices, impeding progress and undermining the realization of their potential benefits(42).

### Data heterogeneity and its challenges

The lack of uniformity in data collection, storage formats, and protocols across different healthcare systems poses a considerable challenge to the widespread adoption of AI in clinical practice. Data standardization is crucial for AI algorithms to effectively analyze and interpret medical information. However, healthcare institutions often employ diverse electronic health record (EHR) systems that vary in their data structures and terminologies. This lack of standardization impedes interoperability and hampers the integration of AI solutions seamlessly. Efforts are needed to establish standardized data formats and protocols, allowing AI systems to operate efficiently across different healthcare settings(43).

### Utilization of AI in Low- and Middle-Income Countries

Implementation of technological advances, including artificial intelligence, in low- and middle-income countries is always a challenge at first because of the high initial investment for capacity-building. The technological and information system infrastructure are varied in LMICs but are often not as established as upper income countries. Furthermore, government support and sheer political will is often lacking posing an added challenge to implementing such efforts in cardiac surgeries in these states.

In spite of this, artificial intelligence advocates have anticipated great gains from implementing artificial intelligence in LMICs. Artificial intelligence offers a unique opportunity to build stronger surgical systems because of its ability to augment clinical judgement (42) and consequently improve diagnostics and therapeutics(44). Such advances may help augment the healthcare system by reducing the needed number of trained specialists and speed up the necessary processes.

### Familiarity and Trust

Familiarity and trust in AI technologies also represent potential barriers to their application in clinical practice. Healthcare professionals may exhibit reluctance or skepticism toward AI, fearing that these technologies may replace their expertise or compromise patient safety. Building trust and familiarity among healthcare providers is crucial for the successful integration of AI in cardiovascular surgery. Transparency in AI algorithms, robust validation studies, and demonstrating tangible benefits can help alleviate concerns and foster acceptance among clinicians(45).

### Risks of Implementing AI

AI algorithms heavily rely on the quality and quantity of input data. If the data used to train these algorithms are incomplete, biased and inaccurate, the AI system could produce unreliable results, potentially leading to incorrect surgical decisions. Safeguards include data validation and data normalization to ensure accuracy and completeness of training data (46).

Deep learning models are often considered black boxes, meaning they provide results without clear explanations of how those results were derived. The lack of interpretability can be problematic especially in critical medical settings like cardiac surgery where clinicians need to understand the rationale behind AI recommendations (46). Some possible measures to overcome this include AI models with built-in explainability features, transparent documentation, healthcare professional training, and regulatory standards.(47)

Physicians may become overly reliant to AI systems, which could lead to medical errors and complacency. Healthcare professionals’ clinical judgment and decision-making skills might diminish in the long run. AI should only be used as a tool to support, rather than replace clinical judgment and expertise. It is vital to maintain a balance between leveraging AI technologies and retaining human expertise in cardiac surgery (48).

### Recent Advances and Future of AI in Cardiac Surgery

The potential use of AI Large Language Models (LLM) such as ChatGPT, is postulated to aid in various aspects of cardiovascular surgery, which may include preoperative planning, intraoperative decision support, and even postoperative care.(49) As a Generative Pre-Trained Transformer (GPT) AI language model, its primary strength lies in its capacity to process large amounts of information and draw out the necessary and most relevant aspects. It is then able to summarize this data and provide concise and contextually appropriate almost human-like responses depending on the specific situation. Intraoperatively, it may potentially be used to provide present-time information about the patient regarding his history and records, or about monitoring his vital signs in the operating room. It may effectively act as a “virtual surgical assistant” that could assist the surgical team. Additionally, it could also help summarize key information relevant specifically to the patient regarding treatment, symptoms, follow-up protocols, and other updates postoperatively. Thus, ChatGPT may aid in delivering more personalized and patient-specific care.(49,50)

A recent study done by Ouyang et al. focused on the development of a deep learning algorithm called “PreOpNet”, which primarily utilized 12-lead Electrocardiogram (ECG) waveform signals, along with other relevant clinical data to help predict postoperative mortality. It was trained using a single preoperative 12-lead ECG result taken within 30 days preoperatively, and appears to be comparable, and in some contexts may perform better than other risk calculators such as the Revised Cardiac Risk Index (RCRI).(51)

Apart from patient care, AI may help in training educating the next generation of cardiac surgeons. Using real and simulated surgical videos and other relevant data as inputs, certain algorithms can be trained on the movements of surgeons to determine which aspects of surgical skills would possibly lead to better surgical performance. These AI algorithms leverage movement tracking and perform kinematic and pose analyses to find which specific details of a surgeon’s movements would be optimal. As a result, the algorithms would theoretically be able to learn which factors are considered surgical expertise, and ideally should be kept in mind to have a better overall surgical performance. With this information, both expert and trainee surgeons would benefit by studying the analyses produced by the novel AI algorithms in order to ideally improve their own respective skillsets. (52)

Recently, augmented reality (AR) or virtual reality (VR) in combination with AI has also garnered attention over the years. Both AR and VR are able to create a real world simulation that allows direct interaction of users with the specified environment. AR creates digital simulations that are directly integrated and projected to the user’s physical environment, while VR creates a digital simulation that is absolutely separate from the user’s environment.(53) Through a dynamic 3-D view of the anatomy, the team would have better insight of the individual’s complex and unique anatomical structures, providing valuable information for better navigation during the surgery. With the use of AI, the technology would be able to process and leverage patient records and imaging files (e.g. preoperative CT scans) in order to improve the rendered AR or VR 3D simulation. The potential clinical feasibility of a combined VR/AR and AI approach has also been demonstrated in fairly recent studies done by Sadeghi et al. and Bakhuis et al. which showcased the use of a combined VR and AI strategy in providing better visualization of their patient’s’ pulmonary anatomy. (54,55)

### Limitations

The quality of research in AI implementation in cardiac surgery needs to be improved. Our review lacks randomized clinical trials (RCTs) and only included cohort studies. In addition, most of the studies acquired clinical data through database registry. Consequently, additional prospective RCTs are necessary to improve the generalizability of results.

The application of AI is dependent on robust data, availability of computational ML techniques appropriate for the complex data, and validation of its clinical application. Because the availability of resources is crucial in its implementation to real-life settings, the vast majority of the included studies were done in developed countries.

## CONCLUSION

While the application of artificial intelligence in cardiac surgery has greatly progressed in recent years, more highly powered studies need to be done to assess challenges and to ensure accuracy and safety for use in clinical practice. AI may be better leveraged for screening and diagnosis to facilitate timely treatment of cardiovascular diseases both in high and low resource settings.

In general, although AI implementations in cardiac surgery are in a continuous process of ongoing development, they have shown considerable potential in improving surgical outcomes, enhancing patient care, and optimizing various clinical processes. Future endeavors in research and development should primarily focus on refining AI algorithms, validating their clinical utility through rigorous studies, and eventually integrating them into routine clinical practice. Apart from clinical applications, surgical education for trainees and patient education may also be explored for further applications of this disruptive technology.

## Data Availability

All relevant data are within the manuscript and its Supporting Information files.

## Acknowledgemen

None

---

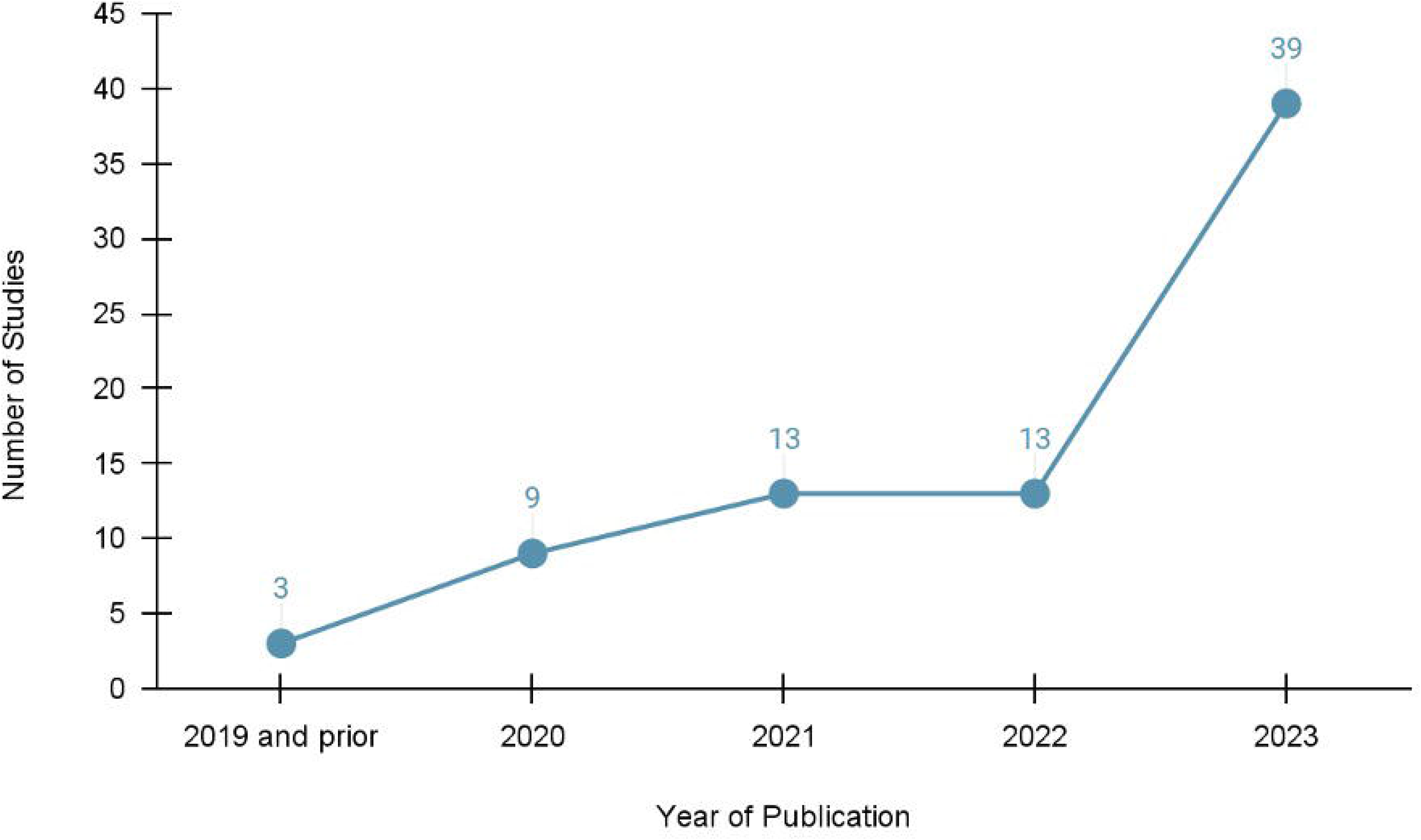

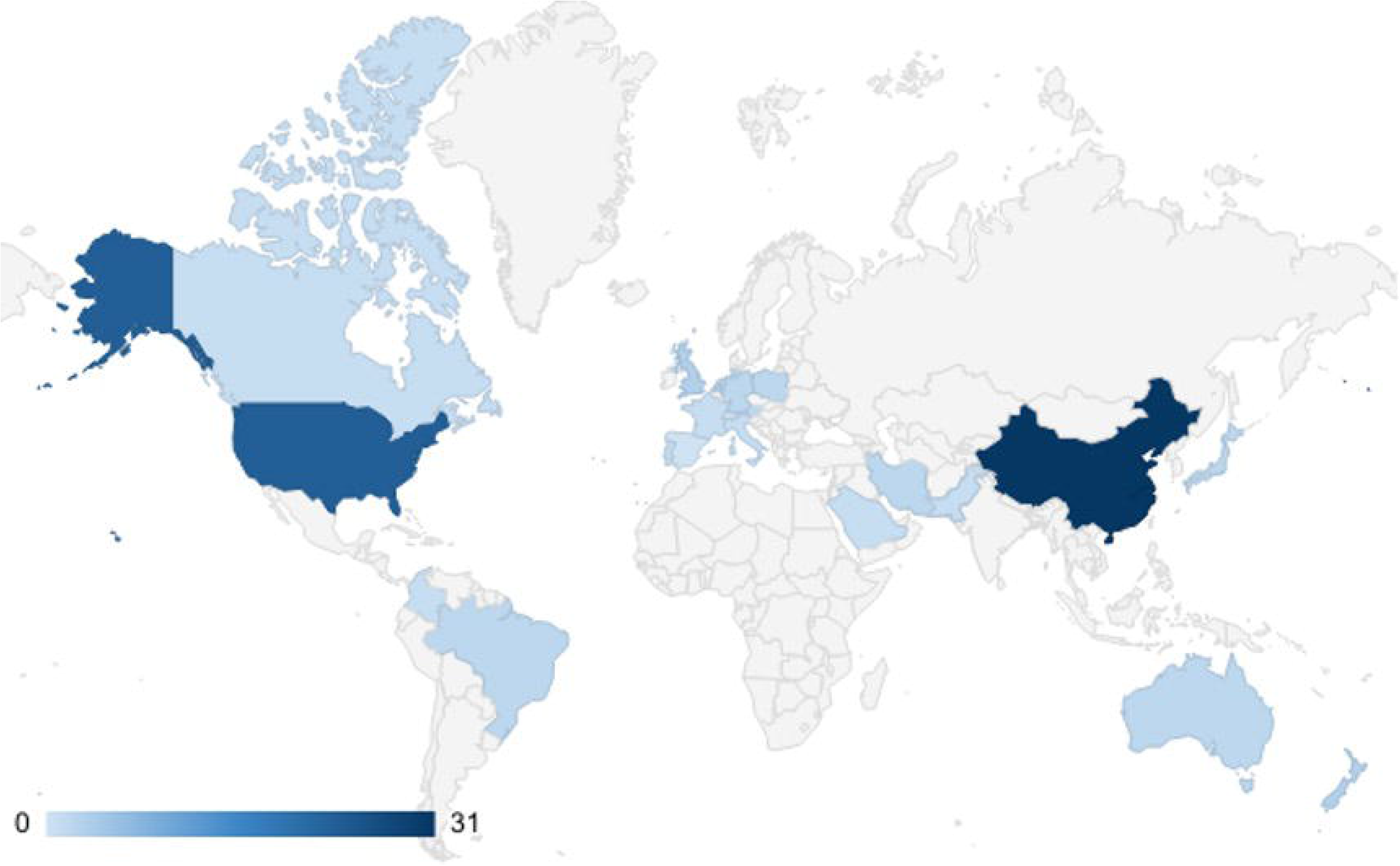

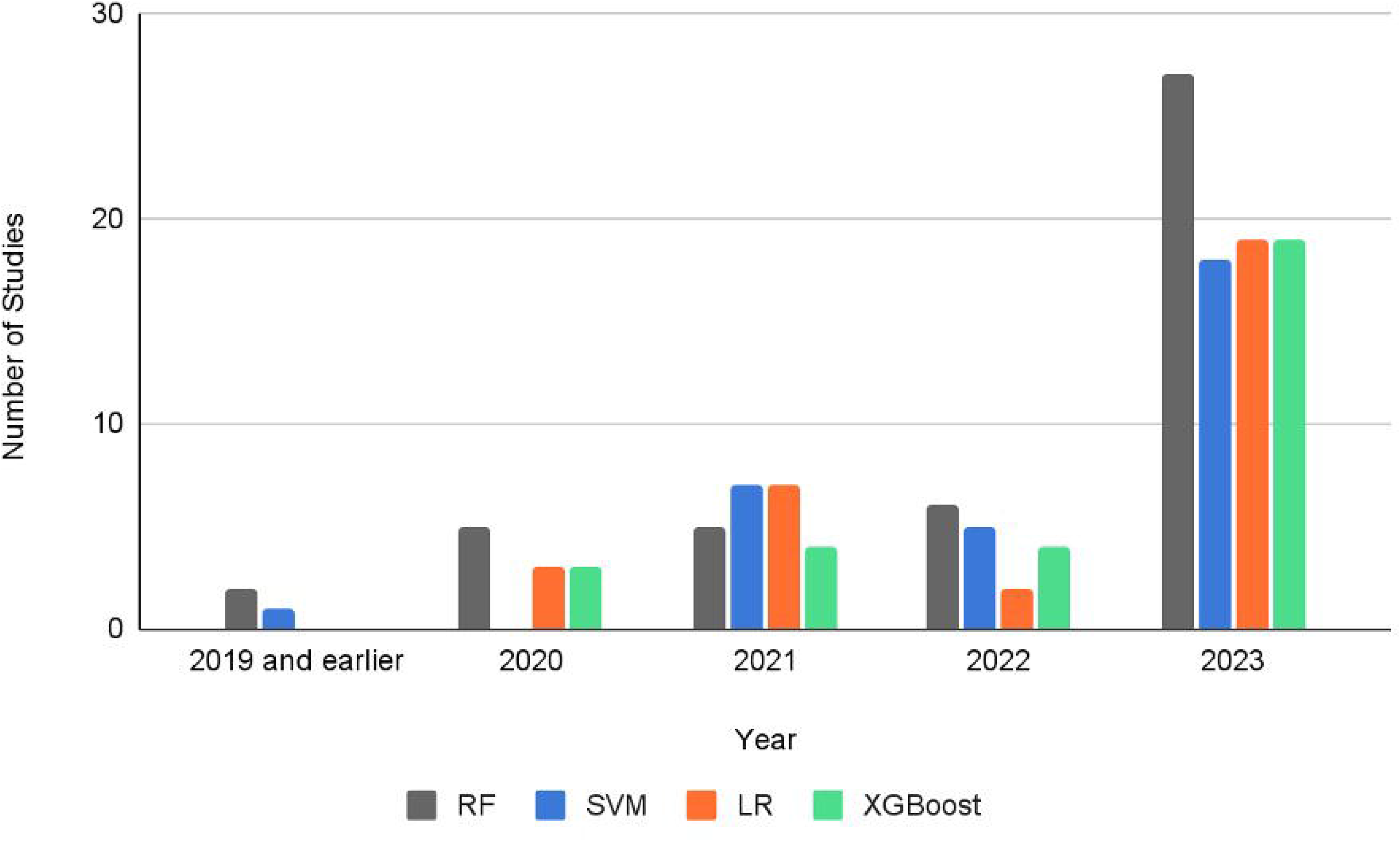

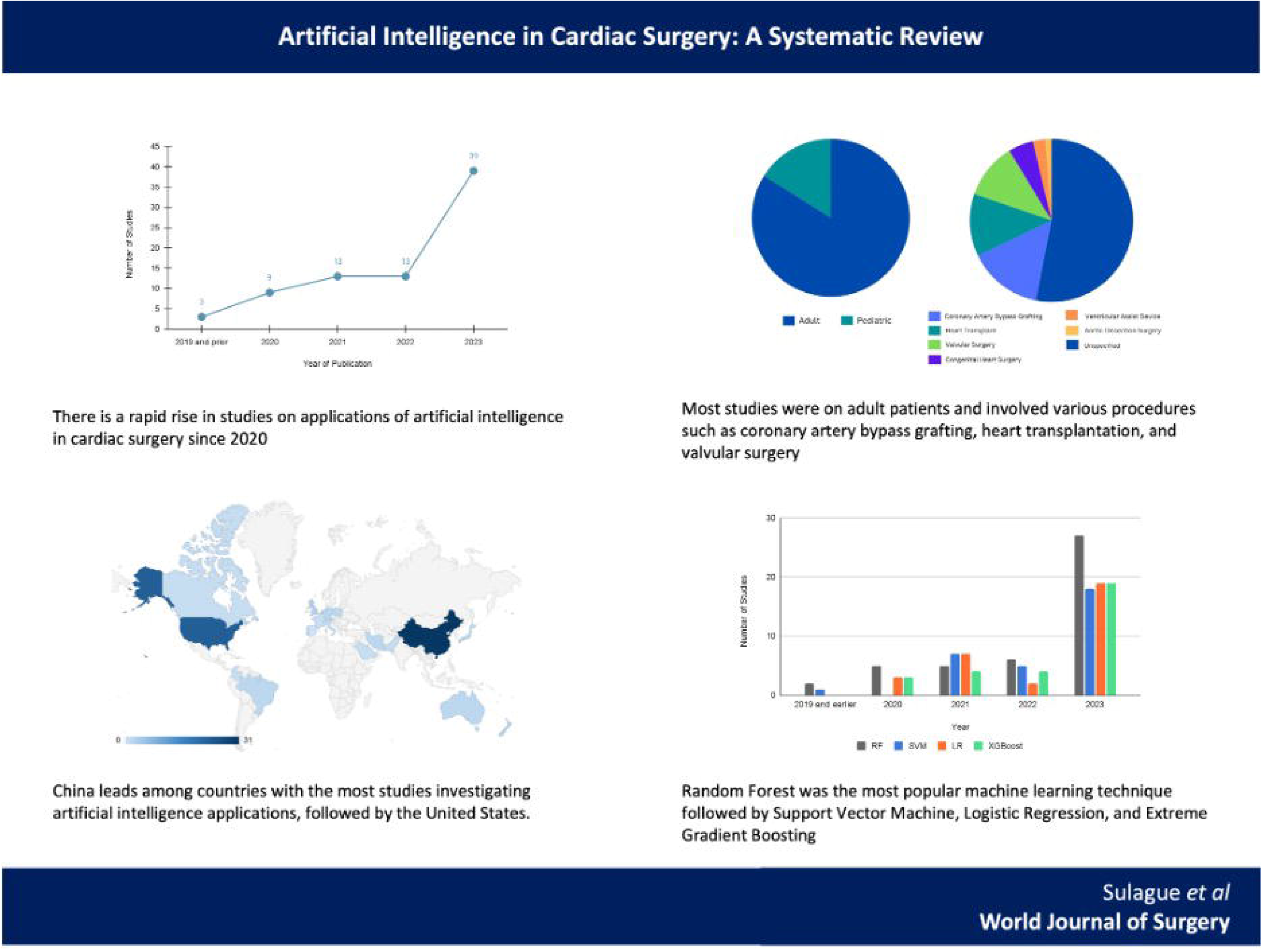

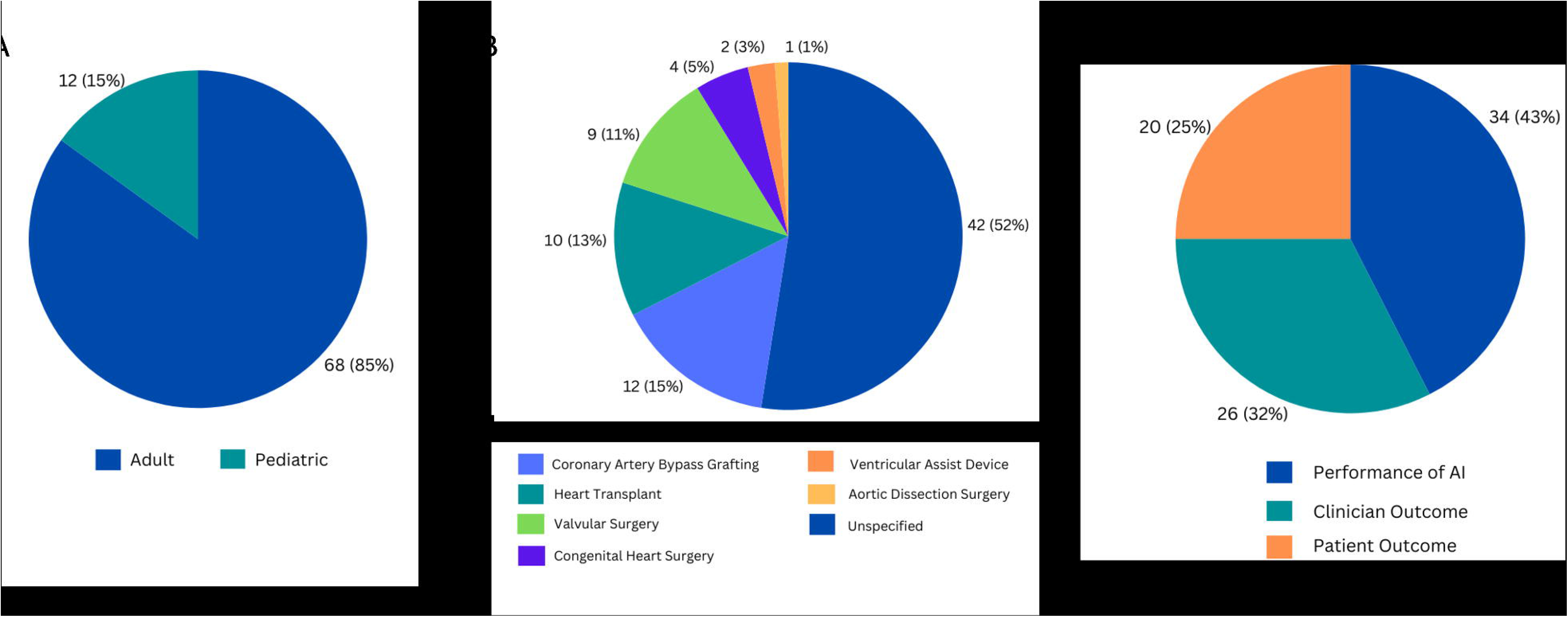

