## Supplemental Table 1 for "Artificial Intelligence in Cardiac Surgery: A Systematic Review"

**Supplement Table 1.** AI Application Characteristics of included studies.

| **Author & Year** | **AI applications** | **AI techniques** | **Clinical Tasks** | **Disease domains and conditions** |
| --- | --- | --- | --- | --- |
| Abdurrab et al., 2024 | Bayesian Models (BM) (Simple and Hierarchical), Stochastic Gradient Descent Regression, Huber Regression, Extreme Gradient Boost (XGBoost), Random Forests (RF), Lasso Regression, Ridge Regression, Support Vector Regression (SVR), Multiple Linear Regression | Evaluate the use of Bayesian regression mod-  els (simple and hierarchical) for Length of Stay (LoS) prediction for cardiac patients (based on their pre-  operative clinical features only) who had undergone cardiac bypass surgery (CABG) | RA | Coronary Artery Bypass Graft (CABG) |
| Agasthi et al, 2020 | Gradient Boosting Machine learning (GBM) | Develop a risk prediction model using a machine learning to predict survival and graft failure (GF)  5 years after orthotopic heart transplant (OHT) | RA | Heart Transplantation (HT) |
| Allyn et al, 2017 | Random Forests,  Gradient Boosting Machine,  Support Vector Machine,  Naïve Bayes Model | compare a machine learning-based model with EuroSCORE II to predict mortality after elective cardiac surgery, using ROC and decision curve analysis. | RA | Cardiac Surgery |
| Alshakhs et al, 2020 | Naïve Bayes,  Decision Tree,  Logistic Regression,  K Nearest Neighbor,  Random Forest | develop and evaluate a model to predict postoperative length of stay (PLoS) for iCABG patients | RA | Coronary Artery Bypass Graft (CABG) |
| Aranda-Michel et al, 2021 | Logistic regression models  imbalanced random forest classifier  Cox regression models,  random forest survival models | learning to evaluate the relationship between MELD score and outcomes of cardiac surgery | RA | Cardiac Surgery |
| Ayers et al, 2021 | deep neural network,  logistic regression,  AdaBoost, and  random forest. | to improve prediction of survival after orthotopic heart transplantation (OHT) | RA | Heart Transplantation (HT) |
| Behnoush et al., 2023 | Logistic Regression (LR), Random Forest (RF), Artificial Neural Network (ANN), Naïve Bayes (NB), Extreme Gradient Boost (XGBoost) | Design and evaluate five ML models to predict 1‐year mortality among hypertensive  patients who underwent CABG | RA | Coronary Artery Bypass Graft (CABG) |
| Betts et al, 2023 | Logistic Regression with Elastic Net regularization (ENR), Gradient Boosting trees (GBT) and artificial neural networks (ANNs) | Construct a risk  model to predict 30-day mortality following pediatric cardiac  surgery specific to patient populations in ANZ by comparing 2  cutting-edge ML approaches with a traditional GLM | RA | Pediatric Cardiac  surgery |
| Bodenhofer et al, 2021 | random forest  neural network  support vector machine | Predict outcome of heart valve surgery with high precision | RA | Heart Valve Surgery |
| Boucek et al, 2023 | Logistic Regression (LR), Random Forest (RF), Adaptive Boosting (AdaBoost), XGBoost, Support Vector Machine (SVM) | Develop a comprehensive ventricular assist device  (VAD) risk assessment tool with the goal of predicting mortality  risk for children being considered for VAD support | RA | Pediatric Ventricular Assist device (VAD) |
| Chang Junior et al, 2020 | Multilayer Perceptron (MLP)  Random Forest (RF)  Extra Trees (ET)  Stochastic Gradient Boosting (SGB)  Ada Boost Classification (ABC)  Bag Decision Trees (BDT) | generate a predictive model calculator adapted to the regional reality focused on individual mortality prediction among patients with congenital heart disease undergoing cardiac surgery. | RA | Congenital Heart Disease |
| Dai et al., 2023 | Extreme Gradient Boost (XGBoost), Logistic Regression (LR), Random Forest (RF) Classifier, Support Vector Machine (SVM) | Develop  machine learning (ML) models combined with intraoperative medicine and blood pressure  time-series data to improve the accuracy of early prediction for postoperative AKI risk after the repair of  Type A acute aortic dissection (TA-AAD) | RA | Acute Kidney Injury |
| Dimri et al., 2023 | Gradient Boosting Machine (GBM), Random Forest (RF), Decision Tree (DT), | Compared model fit of tree-based machine  learning to Cox proportional hazards modeling and  aimed to use the best-performing model to determine the key  predictor variables for long-term mortality after cardiac surgery | RA | Cardiac Surgery |
| Dryden et al., 2023 | Random Forest (RF) Regression, Penalized Linear Regression, K-Nearest Neighbors (KNN), Gradient Boosting, Multivariate Adaptive Regression Splines (MARS) | Developed and validated algorithms for predicting  the warfarin dose required to attain a therapeutic INR at the  time of discharge among patients who have undergone cardiac  surgery | DST | Cardiac Surgery |
| Fan et al, 2022 | random forest  neural network  support vector machine  gradient boosting machine | to predict postoperative mortality in patients following cardiac surgery | RA | Cardiac Surgery |
| Fan et al., 2023 | Deep Forest (DF) Model, Random Forest (RF) Model, Extreme Gradient Boosting (XGBoost) | Develop two types of models for cardiac surgery-associated acute kidney injury (CSA-AKI)  based on early postoperative biomarkers, applying multivariate  logistic regression method and machine learning (ML) algorithms | RA | Acute Kidney Injury |
| Fernandes et al, 2020 | Logistic Regression,  Random Forests,  Neural Networks,  Support Vector Machines,  Extreme Gradient Boosting (XGB) | incorporating intraoperative risk factors in predicting mortality following cardiac surgery. | RA | Cardiac Surgery |
| Gao et al, 2022 | support vector machines (SVM)  stochastic gradient boosting (SGBT)  extreme gradient boosting (XGBoost)  random forest (RF)  conditional inference random forest (CIRF)  boosted classification trees, Naïve Bayes (NB)  Classification and regression tree (CART) | evaluated the performance of machine learning (ML) methods to predict postoperative major bleeding. | RA | Postoperative Major Bleeding |
| Gao et al., 2023 | Extreme Gradient Boosting (XGBoost), Logistic Regression (LR) | To derive and validate a machine learning (ML) prediction model of acute kidney injury (AKI) that could be used for  AKI surveillance and management to improve clinical outcomes. | RA | Acute Kidney Injury (AKI) |
| Hasimbegovic et al, 2021 | AutoML | to model the complex decision-making process of Heart Teams when treating young patients with severe symptomatic aortic stenosis with either TAVR or iSAVR and to identify the relevant considerations. | RA | Aortic Stenosis |
| Hata et al., 2023 | Extra Trees Classifier Model | to develop an accurate algorithm to predict (postoperative delirium) POD using EEG  data obtained from portable device. | RA | Postoperative delirium (POD) |
| Hayward et al, 2023 | Random Forest (RF) Model, Logistic Regression (LR) Model | Determine the oxygen delivery indexed to body surface area (DO2i) threshold  associated with postoperative AKI in pediatric patients during cardiopulmonary bypass  (CPB), and whether  it remains clinically important in the context of other known independent risk  factors | RA | Acute Kidney Injury (AKI) after |
| He et al, 2022 | Support vector machine (SVM) | to build a statistical model and machine learning model to predict Postoperative atrial fibrillation (POAF) in patients with preoperative sinus rhythm after cardiac surgery using portable long term ECG monitoring. | RA | Postoperative Atrial Fibrillation (POAF) |
| Hong et al., 2023 | Support Vector Machine (SVM), Logistic Regression (LR), Random Forest (RF) Classifier, Extreme Gradient Boosting (XGBoost), Deep Neural Network (DNN) | timely predict the LCOS risk during post-  operative critical care, allowing sufficient time for clinical intervention, which improves patient outcomes and reduces the occupation of medical resources. | RA | Low cardiac output syndrome (LCOS), |
| Hosseininezhad et al, 2021 | Logistic Regression (LR), linear discriminant analysis (LDA), support-vector machine (SVM), K-nearest neighbors(KNN), and multilayer perceptron (MLP). | generate a machine learning (ML)-based model to predict in-hospital mortality after isolated mitral valve replacement(IMVR). | RA | Isolated Mitral Valve Replacement (IMVR) |
| Jia et al., 2023 | Light Gradient Boosting Machine (LightGBM), Support Vector Machine (SVM), Softmax Regression, Random Forest (RF) | to apply a new machine learning (ML) method to establish prediction  models of AKI after CABG. | RA | Acute kidney injury (AKI) |
| Jiang et al, 2021 | eXtreme Gradient Boosting (XGBOOST)  CatBoost, LightGBM, MLP, SVM,  LR, Random Forest, Gradient Boosting Machine, KNN, AdaBoost, and  Naive Bayes | identify critical preoperative and intraoperative variables and predict the risk of several severe complications (myocardial infarction, stroke, renal failure, and hospital mortality) after cardiac valvular surgery. | RA | Perioperative Complications |
| Jiang, J. et al., 2023 | Gradient Boosting Classifier (GBC), Decision Tree (DT), Random Forest (RF), Gaussian Naive Bayes, Multilayer Perceptron | Evaluate whether machine learning algorithms could significantly improve the risk prediction of PO-AKI. | DST | Postoperative acute kidney injury (PO-AKI) |
| Jiang, Z. et al., 2023 | Category Boosting (CatBoost), Extreme Gradient Boosting (XGBoost), Light Gradient Boosting Machine (LightGBM), Random Forest (RF), Support Vector Machine (SVM), Logistic Regression (LR), Adaptive Boosting (AdaBoost), Bootstrapped Aggregation (Bagging), Gradient Boosting Decision Trees (GBDT), Multilayer Perceptron (MLP) | Evaluate the efficacy of the Cox-Maze IV procedure (CMP-IV) in  combination with valve surgery in patients with both atrial fibrillation (AF) and  valvular disease and use machine learning algorithms to identify potential risk  factors of AF recurrence. | RA | atrial fibrillation (AF) and  valvular disease |
| Just et al., 2024 | Convolutional Neural Network | Examine the preoperative computed tomography (CT) body composition as a predictor of the postoperative outcome  in advanced HF patients, who receive LVAD implantations. | DST | Heart failure (HF) |
| Kampaktsis et al, 2021 | Adaboost,  Logistic Regression,  Decision Tree,  Support Vector Machine,  K-nearest neighbor models | to develop and validate machine learning (ML) models to increase the predictive accuracy of mortality after heart transplantation (HT). | RA | Heart Transplantation (HT) |
| Kampaktsis et al, 2022 | CatBoost ML model | develop and validate an ML model for the prediction of mortality after heart transplantation (HT) in adults with congenital heart disease (ACHD). | RA | Heart Transplantation (HT) |
| Karri et al, 2021 | Random Forest Classifier (RF),  Decision Tree Classifier (DT),  Logistic Regression (LR),  K Neighbors classifier (KNN),  Support Vector Machine (SVM),  Gradient Boosted Machine (GBM) | compared the performance of machine learning (ML) to the to the established gold standard scoring tool (POAF Score) in predicting postoperative atrial fibrillation (POAF) during intensive care unit (ICU) admission after cardiac  surgery. | RA | Postoperative Atrial Fibrillation (POAF) |
| Kilic et al, 2020 | Extreme gradient boosting (XGBoost) | estimating operative mortality risk in cardiac surgery | RA | Cardiac Surgery |
| Kim et al, 2022 | Dual-tree complex wavelet packet transform (DTCWPT)18 | that predict the occurrence of several life-threatening complications up to 4 hours prior to the event. | RA | Perioperative Complications |
| Kobayashi et al., 2023 | Recurrent Neural Network, Linear Regression, Random Forest (RF), Artificial Neural Network, Transformer | Use several machine learning and  deep learning approaches to predict maximum blood lactate  concentrations in patients up to 24h after cardiac surgery. | RA | Cardiac Surgery |
| Kong et al, 2023 | Extreme gradient boosting (XGB), Logistic regression (LR), Light gradient boosting machine (LGBM),  GaussianNB (GNB), Multilayer Perceptron (MLP), Support Vector Machine (SVM) | Develop a prediction model that can be used to accurately predict AKI through machine learning | RA | Acute Renal Injury (AKI) |
| Lee et al, 2013 | GenAlgs | To develop a customized short LOS (<6 days) prediction model for geriatric patients receiving cardiac surgery, | RA | Cardiac Surgery |
| Li et al, 2020 | Bayesian networks (BNs) | To predict the individual risk of CSA-AKI occurrence. | RA | Acute Kidney Injury |
| Li et al, 2022 | Logistic regression with L2 regularization | to predict post-transplant AKI stage 3 based on preoperative and perioperative features. | RA | Acute Kidney Injury |
| Li, Qian et al., 2023 | CatBoost Classifier (CAT), Logistic Regression (LR), Support Vector Machine (SVM), KNeighborsClassifier (KNN) Naive Bayes (BAY), Decision Tree (DT), Random Forest (RF), Gradient Boosting Classifier (GB), XGBoosting Classifier (XGB), Light Gradient Boosting Machine (LGBM), AdaBoost Classifier, Extra Tree Classifier | Develop and validate predictive models based on  a multicenter randomized control trial (RCT) through ML and traditional LR methods. | RA | Acute Kidney Injury |
| Li, Qiuying et al., 2023 | Random Forest (RF) Classifier, Logistic Regression (LR), Support Vector Machine (SVM) Classifier, K-Nearest Neighbors (KNN) Classifier, Gaussian Naive Bayes, Gradient Boosting Decision Tree, Perceptron | To develop and validate a machine learning-based prediction model for postoperative delirium (POD) in cardiac valve  surgery patients. | RA | Postoperative Delirium (POD) |
| Linse et al., 2023 | Artificial Neural Network (ANN) (specifically, Multilayer Perceptron (MLP)) | develop and validate a risk model for  prediction of short-term mortality due to PGF after heart transplantation using the ISHLT Heart  Transplant Registry. | RA | Primary graft failure (PGF) |
| Lo et al, 2021 | Optimizable KNN (k-nearest neighbor classifier),  Optimizable SVM (support vector machine classifier) | to develop Vi.Ki.E.-based SML models to provide  an important decision-making tool supporting the medical team during open-chest surgery. | RA | Open-Chest Surgery |
| Lu et al., 2023 | Support Vector Machine (SVM), Gradient Boosting Decision Tree (GBDT) | develop the predictive models for AF after cardiac surgery by using machine learning approaches, including the gradient-boosting decision  tree (GBDT) and the support-vector machine (SVM), then  compared their performance with the classic LR model. | RA | Postoperative atrial fibrillation (POAF) |
| Luo et al, 2021 | Sun Yat-sen University Prediction Model for Infective Endocarditis | construct an accurate and easy-to-use prediction model to identify patients at high risk of early mortality after surgery for infective endocarditis. | RA | Infective Endocarditis |
| Mathis et al, 2022 | random forest model | early detection of post-operative deterioration among patients undergoing cardiac surgical procedures. | DST | Post-Operative Deterioration |
| Mazhar et al., 2023 | Bayesian Networks |  |  |  |
| Miller et al, 2019 | Artificial Neural Network (ANN)  Classification and Regression Tree (CART)  Random Forest (RF) | evaluated the utility of 3 ML algorithms for predicting mortality after  pediatric HTx | RA | Heart Transplantation (HT) |
| Molina et al, 2022 | Logistic regression models,  Naive Bayes, Multilayer perceptron,  Support vector machine (SVM),  Random forest,  Gradient boosting (boosted trees) | to predict cardiac surgical operative mortality. | RA | Cardiac Surgery |
| Nowakowska et al., 2023 | Random Forest (RF) Classifier, Gradient Boosted Tree (GBT), Adaptive Boosting (AdaBoost), Extreme Gradient Boosting (XGBoost) | Predict both depression and postoperative delirium among  patients who underwent CABG. | RA | Postoperative Delirium Depression |
| Nowicka-Sauer et al., 2023 | Classification and Regression Tree (CART), K-Nearest Neighbor (KNN), Support Vector Machine (SVM), Multilayer Perceptron (MLP), Naive Bayes Classifier | Construct a model of the risk factors of depression in patients following cardiac surgery, with the use of machine learning. | RA | Depression |
| Park et al, 2022 | Logistic Regression (LR) model, Adaptive  Boosted (AdaBoosted) Trees, Bootstrap Aggregating (Bagged)  Trees, Subspace discriminant, Subspace K-nearest neighbor  (KNN), Random under-sampling Boosted (RUSBoosted) Trees, XGboost | to analyze and predict outcomes after open-heart surgery. | RA | Open-Heart Surgery |
| Parise et al., 2024 | Random Forest (RF), Multivariate Adaptive Regression Splines (MARS), Neural Network, Support Vector Machine (SVM) | to get an effective machine learning (ML) prediction model of  new-onset postoperative atrial fibrillation (POAF) following coronary artery bypass grafting (CABG)  and to highlight the most relevant clinical factors. | RA | Postoperative atrial fibrillation (POAF) |
| Raghu et al, 2022 | deep learning model (CXR-CTSurgery) | develop a model that estimate postoperative mortality risk based on a preoperative chest radiograph for cardiac surgeries | RA | Cardiac Surgery |
| Santos R. et al., 2023 | Random Forest (RF), Support Vector Machine (SVM), Naive-Bayes (NB), Decision  Tree (DT), Light Gradient Boosting Machine (LGBM) | Clinical Decision Support System based on Machine Learning to estimate the risk of severe  complications within 90 days following cardiothoracic surgery discharge. | RA | Post-Discharge Complications |
| Shao et al., 2023 | K-Nearest neighbor, Logistic Regression, Decision Tree (DT),  Random Forest (RF), Support Vector Machine (SVM), Neural Network | Develop models for predicting CSA-  AKI | RA | Acute Kidney Injury |
| Shou et al, 2022 | extreme gradient boosting (XGBoost) | prediction of post-transplant mortality in patients bridged to heart transplantation with temporary mechanical circulatory support (tMCS) | RA | Heart Transplantation (HT) |
| Simons et al., 2023 | Linear Regression, Random Forest (RF), Extreme Gradient Boosting (XGBoost) | to evaluate the performance of the  current surgery duration model used in clinical practices, (ii) to  develop and validate an enhanced predictive model and (iii) to get  insight into which patient and surgery characteristics are key  features in the model development. | RA | Cardio-thoracic surgery duration |
| Sinha et al., 2023 | Extreme Gradient Boosting (XGBoost), Random Forest (RF) | To perform a systematic comparison of in-hospital mortality risk prediction post-cardiac surgery, between the predominant scoring system—European System for Cardiac Operative Risk Evaluation (EuroSCORE) II, logistic regression (LR) retrained on the same vari-  ables and alternative machine learning techniques (ML)—random forest (RF), neural networks (NN), XGBoost and weighted support vector  machine. | RA | Mortality risk prediction post-surgery |
| Sughimoto et al, 2020 | Hypertuned Random Forest, Random Forest Regressor, AdaBoost Regressor, Hypertuned AdaBoost, Decision  Tree, and Hypertuned Decision tree. | Prediction of blood lactate levels in pediatric ICU patients using machine learning applied to arterial waveforms  and perioperative characteristics | DST | Blood Lactate Levels (as marker of hemodynamic stability/instability) |
| Tong et al., 2023 | Random Forest Classifier (RFC), Extreme Gradient Boosting (XGBoost), Logistic Regression (LRC), Light Gradient Boosting Machine (LGBM), Adaptive Boosting (AdaBoost), and K-nearest neighbor (KNN) | Use machine learning models to construct models to predict acute kidney injury after extracorporeal cardiac surgery (CSA-AKI) and screen for  the best predictive model | RA | Acute Kidney Injury after extracorporeal cardiac surgery (CSA-AKI) |
| Tong et al, 2024 | Light Gradient Boosting Machine (LightGBM), Logistic Regression (LR), Support Vector Machine (SVM), Random Forest (RF), CatBoost | Evaluate the performance of five machine learning algorithms for predicting four major adverse postoperative outcomes (APOs) after pediatric congenital heart surgery and their clinically meaningful  model interpretations and translational impact | RA | Congenital Heart Surgeru |
| Wang et al, 2022 | Gaussian Process (GP) regression ML algorithm  hybrid machine learning (ML) framework:  Random Forest (RF) algorithm,  Gaussian process (GP) regression model, compared the performance of GP classification to neural networks, XGBoost, random forest, decision tree | predicting red blood cell (RBC) transfusion requirements in cardiothoracic (CT) surgery | RA | Red Blood Cell Transfusion |
| Weiss et al., 2023 | Extreme Gradient Boosting (XGBoost),  Random Forest (RF), Logistic Regression (LR), Support  Vector Machine (SVM) | Develop a rigorous machine learning framework applied to routinely collected, multi-modal EHR data from a large, all-comers cardiac surgery patient cohort could to create a personalized,  institution- specific risk prediction model for mortality. | RA | Post-operative Mortality |
| Williamson et al, 2023 | Logistic  Regression with Ridge Penalization, K-nearest neighbors, Support Vector Machine (SVM), Decision tree, Random Forest  models | Trained models to classify the risk of postoperative infections using logistic regression and several  machine learning methods | RA | Postoperative Infections |
| Wisotzkey et al, 2023 | Cox Regression, Gradient Boost-  ing, Axis-based random survival forests, Oblique random survival forests | Determine if machine learning could improve 1-year risk assessment using the Pediatric Heart Transplant Society database | RA | Heart Transplant (HT) |
| Wu et al.,  2023 | Extreme Gradient Boosting (XGBoost),  Logistic  Regression, Simple Decision Tree, Random Forest (RF), Support  Vector Machine (SVM) | Establish a predictive model  for preoperative in-hospital mortality of patients with  acute aortic dissection (AD) by using machine learning  techniques | DST | Acute Aortic Dissection |
| Xue et al, 2022 | Multivariate logistic regression (MLR),  Support vector machine learning (SVM),  Random forest model (RFM),  Multilayer perceptron (MLP) | Establishing and validating a machine learning-based quantitative method for noninvasive hippocampal assessment through preoperative cranial computed tomography (CT) instead of magnetic resonance imaging for the early detection of AKI-related hippocampal damage and prompt clinical intervention | DST | AKI - related Hippocampal Damage |
| Yan et al., 2023 | Lasso Logistics Regression (LLR), Random Forest (RF), and  Extreme Gradient Boosting (XGboost) | Develop prediction models for AKI after valvular cardiac  surgery in the Chinese population | RA | Postoperative Acute Kidney Injury (AKI) |
| Zea-Vera et al, 2021 | Extreme Gradient Boosting (XGBoost) | Develop and validate a dynamic machine learning model to predict CABG outcomes at clinically relevant pre- and postoperative time points | RA | Coronary Artery Bypass Graft (CABG) |
| Zeng et al, 2021 | XGBoost | Develop an interpretable machine-learning based model that integrates patient demographics,  surgery-specific features and intraoperative blood pressure data for accurately predicting  complications after pediatric congenital heart surgery | RA | Postoperative Complications |
| Zeng et al, 2023 | Support Vector Machine (SVM),  Logistic Regression (LR),Long short-term memory (LSTM) RNN, Gate Recurrent Units (GRU) RNN, Dipole, RETAIN, Proposed Time-Aware Attention RNN | Develop and validate a model for predicting  postoperative Acute Kidney Injury (AKI) that operates sequentially over individual time-series clinical data | DST | Postoperative Acute Kidney Injury (AKI) |
| Zeng et al., 2023 | Random Forest (RF), Logistic Regression  (LR), Decision Tree (DT), Extreme Gradient Boosting (XGBoost) | Establish a risk prediction model including intraoperative features set as the primary objective variables, with Off-pump coronary artery bypass grafting-associated acute kidney injury (OPCAB-AKI) set as the sole outcome. | RA | Off-pump coronary artery bypass grafting-associated acute kidney injury (OPCAB-AKI) |
| Zhang et al., 2023 | Random forest (RF), Extreme  Gradient Boosting (XGBoost), Support Vector Machine (SVM), Gradient Boosting  Decision Tree (GBDT), AdaBoost, Naive Bayesian (NB), Logistic Regression  (LogicR), Neural Networks (Nnet), Artificial Neural Network (ANN) | Used machine  learning methods to identify critical perioperative infection- related variables after  mitral valve surgery and construct a prediction model | RA | Perioperative Infection |
| Zheng et al., 2023 | Random Forest (RF), Support Vector Machine (SVM) with  Radial Basis Function Kernel, Extreme Gradient Boosting  (XGBoost) | Use regression and machine learning models to identify a combination  of cardiac magnetic resonance (CMR) LV remodeling and function parameters that predict LVEF < 50% after  mitral valve surgery | RA | Mitral Valve Surgery |
| Zhong et al, 2020 | Logistic Regression (LR)  Artificial Neural Network (ANN)  Random Forest (RF)  Extreme Gradient boosting (XGBoost) | Build up multiple machine learning models to predict 30-days mortality, and 3 complications including septic shock, thrombocytopenia, and liver dysfunction after open-heart surgery. | RA | Perioperative Complications |
| Zhou et al, 2021 | Logistic regression (LR),  Support Vector Machines (SVM)  Random Forest (RF)  Extreme gradient boosting (XGBoost),  Adaptive boosting (AdaBoost),  Gradient boosting machine (GBM)  Artificial neural network (ANN) | Establish a risk-prediction model for assessing prognosis of HTx using machine-learning approach. | RA | Heart Transplantation (HT) |
| Zhu et al., 2023 | Support Vector Machine (SVM),  by Logistic regression  (LR),  Complement Naive Bayes (CNB) model | Compare different machine learning algorithms to predict individual risk of POAF after valve surgery and to identify the most influential preoperative and intraoperative variables. | RA | Postoperative Atrial Fibrillation  (POAF) |
| Zurn et al, 2023 | Logistic Regression (LR) Model | Improve postoperative risk and survival assessment in congenital heart surgery by developing a machine-learning model based on readily available peri- and postoperative parameters | RA | Congenital Heart Surgery |

**RA - Risk Analysis*

**DST - Disease Screening or Triage*
